# Evaluation of crowdsourced mortality prediction models as a framework for assessing AI in medicine

**DOI:** 10.1101/2021.01.18.21250072

**Authors:** Timothy Bergquist, Thomas Schaffter, Yao Yan, Thomas Yu, Justin Prosser, Jifan Gao, Guanhua Chen, Łukasz Charzewski, Zofia Nawalany, Ivan Brugere, Renata Retkute, Alidivinas Prusokas, Augustinas Prusokas, Yonghwa Choi, Sanghoon Lee, Junseok Choe, Inggeol Lee, Sunkyu Kim, Jaewoo Kang, Patient Mortality Prediction DREAM Challenge Consortium, Sean D. Mooney, Justin Guinney

## Abstract

Applications of machine learning in healthcare are of high interest and have the potential to significantly improve patient care. Yet, the real-world accuracy and performance of these models on different patient subpopulations remains unclear. To address these important questions, we hosted a community challenge to evaluate different methods that predict healthcare outcomes. To overcome patient privacy concerns, we employed a Model-to-Data approach, allowing citizen scientists and researchers to train and evaluate machine learning models on private health data without direct access to that data. We focused on the prediction of all-cause mortality as the community challenge question. In total, we had 345 registered participants, coalescing into 25 independent teams, spread over 3 continents and 10 countries. The top performing team achieved a final area under the receiver operator curve of 0.947 (95% CI 0.942, 0.951) and an area under the precision-recall curve of 0.487 (95% CI 0.458, 0.499) on patients prospectively collected over a one year observation of a large health system. Post-hoc analysis after the challenge revealed that models differ in accuracy on subpopulations, delineated by race or gender, even when they are trained on the same data and have similar accuracy on the population. This is the largest community challenge focused on the evaluation of state-of-the-art machine learning methods in a healthcare system performed to date, revealing both opportunities and pitfalls of clinical AI.

## Introduction

Applications of machine learning applied to patient data are undergoing wide development and implementation in healthcare scenarios ^1,2^. The performance of these methods as they are used in the clinic - and their associated impact on patient outcomes - are not well understood. An important risk in the design and implementation of machine learning algorithms is the self-assessment bias, where the implementer and evaluator are the same person or team, which can result in overfitting and poor generalization ^3^. At the same time, health systems and journals are inundated with new methods that overwhelm the ability of healthcare providers to assess effective solutions. This is further exacerbated by varying business practices across healthcare institutions resulting in vastly different data characteristics at each site. At each of these institutions, clinical practice and data collection practices can change over time, in some cases, rendering EHR data obsolete in as little as 3-6 months, as it no longer reflects current data distributions ^4^. Healthcare data can also contain hidden biases that reflect social and institutional disparities within clinical practice ^5^. Risks of biases in medicine have been well documented, and models built using biased data will propagate these biases into practice through model recommendations ^5,6^. Addressing these issues requires a rigorous, unbiased framework that can evaluate algorithm performance using independent honest brokers, assess generalizability over time and across institutions, and report on performance disparities across sub-populations.

Access to electronic healthcare record (EHR) data for AI research, and subsequent evaluation, is complicated by HIPAA regulations, privacy concerns, and business risks. To overcome these barriers to access, we have developed an approach called Model-to-Data (MTD) that delivers analytical models to protected data without sharing the data directly with model developers ^7^. We previously piloted this method on an EHR dataset from the University of Washington and demonstrated the feasibility of accurate model development without the model developer having direct access to the patient data ^8^. This approach has two benefits: (1) it protects patient data while allowing researchers to build machine learning methods and (2) it forces a more standardized and transferable approach to building models allowing the data host to perform rigorous evaluations of submitted models.

We leveraged this platform to implement the *EHR DREAM Challenge: Patient Mortality Prediction* to assess machine learning approaches applied to a clinical data warehouse while protecting patient privacy. DREAM Challenges are crowdsourced, biomedical competitions, where the challenge organizers solicit the broader research community to develop methods to answer a specific set of biomedical questions ^9^, and to assess these methods using hidden, gold standard datasets. Community challenges have proven to be a robust setting for the objective evaluation of prediction models since they remove the researcher from the evaluation process ^10–14^, limiting the self assessment bias ^3^. We focused on the clinical question of predicting all-cause mortality, as the clinical phenotype is clearly defined and complete (University of Washington merges patient records with state death records to minimize missingness) and previous mortality prediction methods have been developed ^15–18^. In this Challenge, we asked participants to predict whether patients would pass away within 180 days of their last visit to the UW medical system based on that patient’s previous medical history. We evaluated models for population level accuracy, longitudinal generalizability - evaluating models on a prospectively collected data set - and demographic generalizability - evaluating model performance across sensitive demographic strata.

## Methods

### The University of Washington clinical data repository

The UW Medicine enterprise data warehouse (EDW) includes patient records from clinical sites within the UW Medicine system, including more than 300 specialty and primary care clinics. The EDW gathers data from more than 60 sources, including laboratory results, microbiology reports, demographic data, diagnosis codes, medications prescribed, and procedures performed. An analytics team at the University of Washington transformed the patient records from 2010 to 2019 into a standardized data format, the Observational Medical Outcomes Partnerships Common Data Model (OMOP CDM v5.0) ^19^. For the EHR DREAM Challenge, we selected all patients who had at least one visit in the UW OMOP repository, which represented 1.3 million patients, 22 million visits, 33 million procedures, 5 million drug exposure records, 48 million condition records, 10 million observations, and 221 million measurements covering approximately 10 years of patient histories.

### Synthetic data

We made synthetic data available to participants for local model development and debugging purposes, and used it in the Challenge platform environment to validate that submitted models could be successfully executed. (Figure 1, Stage 1) Running containers on synthetic data removed the privacy concerns intrinsic in real clinical data allowing us to return logs and error messages to participants without screening. We derived a synthetic dataset from the SynPuf Synthetic OMOP dataset ^20^. Starting with the original SynPuf dataset we adapted it to our challenge by randomly sampling concepts and terms that occurred more than 100 times in the structured EHR data from the University of Washington OMOP repository and then populated the tables of the original SynPuf dataset with these random samples to create a synthetic derivative that more closely represents the UW OMOP data. We also adjusted the size of the data to match the distribution of records across patients resulting in a synthetic dataset that represented 1,264,000 patients with 6300 true positives, 19,945,000 visits, and 189,605,000 measurements (see the supplemental material).

**Figure 1.**
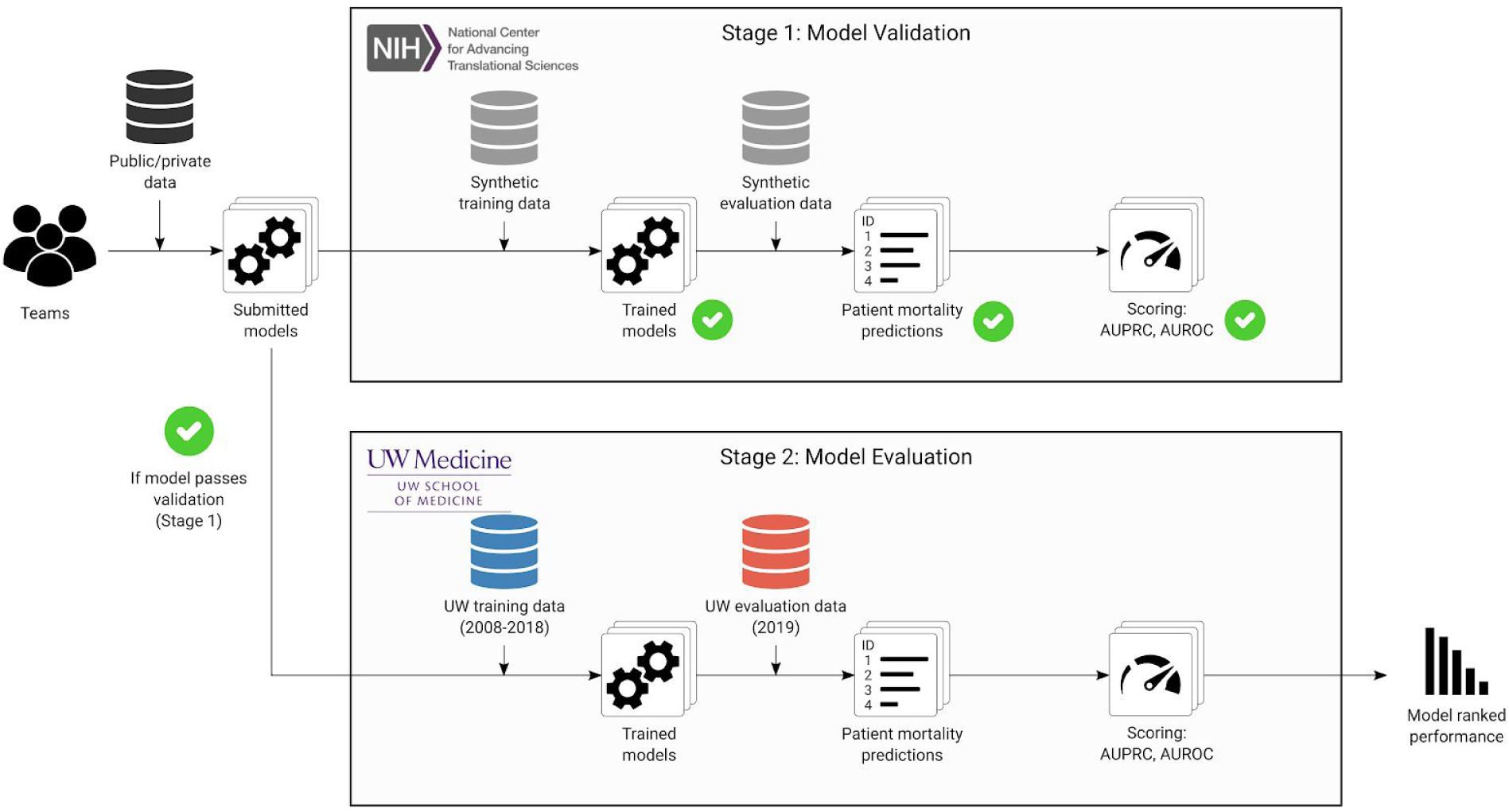
Model-to-data architecture to evaluate the performance of EHR prediction models in the Patient Mortality DREAM Challenge. Models were developed on local environments using synthetic data that resembled the real private EHR data. Docker images were submitted through the Synapse collaboration platform to a submission queue. Images were pulled into the NCATS provided AWS cloud environment and run against a synthetic dataset for technical validation (Stage 1). Once validated, images were pulled into the UW Medicine secure infrastructure and run against the private EHR data. Model predictions were evaluated using Area Under the Receiver Operator Curve (AUROC) and Area Under the Precision Recall Curve (AUPRC) which were returned to participants through Synapse.

### The EHR DREAM Challenge: Patient Mortality Prediction

#### The challenge infrastructure

The EHR DREAM Challenge was developed and run using a “Model to Data” (MTD) approach ^7,8^. This method relies on containerization software (Docker) ^21^, a common data model (OMOP) ^19^, a model intake mechanism (Synapse) ^22^, and a synthetic dataset for low risk technical validation of submitted models (Synpuf) ^20^. Challenge participants were required to submit “containerized” models to be applied to protected data by the Challenge organizers. **At no time during the Challenge did participants have direct access to real patient data and models never had access to patient identifiers**. Participants were allowed to submit *pretrained* models using data they already have, such as their own institution’s clinical data warehouse. The containerized algorithms submitted by participants were able to use a training split of the UW dataset to train a predictive model *de-novo*, or to further optimize a pretrained model. To enable model debugging prior to evaluation on real clinical data, submitted models were first applied to the synthetic data to check for technical compliance (Figure 1, Stage 1: Model Validation). Log files generated by the models in the synthetic data environment were returned to participants for technical debugging purposes. Following successful execution on the synthetic data, the models were pulled into a University of Washington secure environment that was disconnected from the internet where they were trained on the UW OMOP repository. The UW OMOP repository did not contain patient identifiers. The models had no access to the internet during training and evaluation. (Figure 1, Stage 2: Model Evaluation) The trained models were tested on a holdout set and the Area Under the Recall Curve (AUROC) and Area Under the Precision Recall Curve (AUPRC) were returned to participants via the Synapse platform. No logs, model parameters, or other information other than the performance metrics, were returned to participants after models were applied to the UW patient repository (including the final models themselves). Participants were allowed a total of 10 hours to train and test their models in this environment. The models were run on a server environment with access to 70 GB of RAM, 32 2.3 GHz CPU cores and no GPUs during this process. Model predictions were never linked to identified patient records.

#### Challenge Question

For this challenge, we asked participants to predict the 180-day all-cause mortality from the last patient visit at UW Medicine. True positives were defined as patients who had a death record in the first 180 days of their last visit record and true negatives were defined as patients who either had a death record more than 180 days from their last visit, or who did not have a death record. Death records were derived from the medical record and Washington State death records.

#### Timeline

The EHR DREAM Challenge lasted from September 9, 2019 to February 23, 2020 and was conducted in three phases: the open phase, the leaderboard phase, and the validation phase. During the open phase, participants could submit models for technical validation using only synthetic data in the Challenge cloud environment (Figure 1, Stage 1). During the leaderboard phase, models that were technically validated against the synthetic data were applied to the leaderboard training data and evaluated against the leaderboard validation data (Table 1). This phase was designed to give participants the opportunity to build and tune their models, receiving model performance metrics on the leaderboard validation data set. We carried out this phase in 3 rounds, where each team was allowed 3 successful submissions per round. During the open and leaderboard phases, new data accumulated in the UW EHR. We gathered this data (called the “holdout test data”) which represented patients who visited UW medical facilities during 2019. During the validation stage, models were trained on the leaderboard training data (excluding the patients transitioned to the holdout test data) and were tested on this new prospectively-collected data. See supplementary materials for details on the timeline and data generation processes.

**Table 1.**
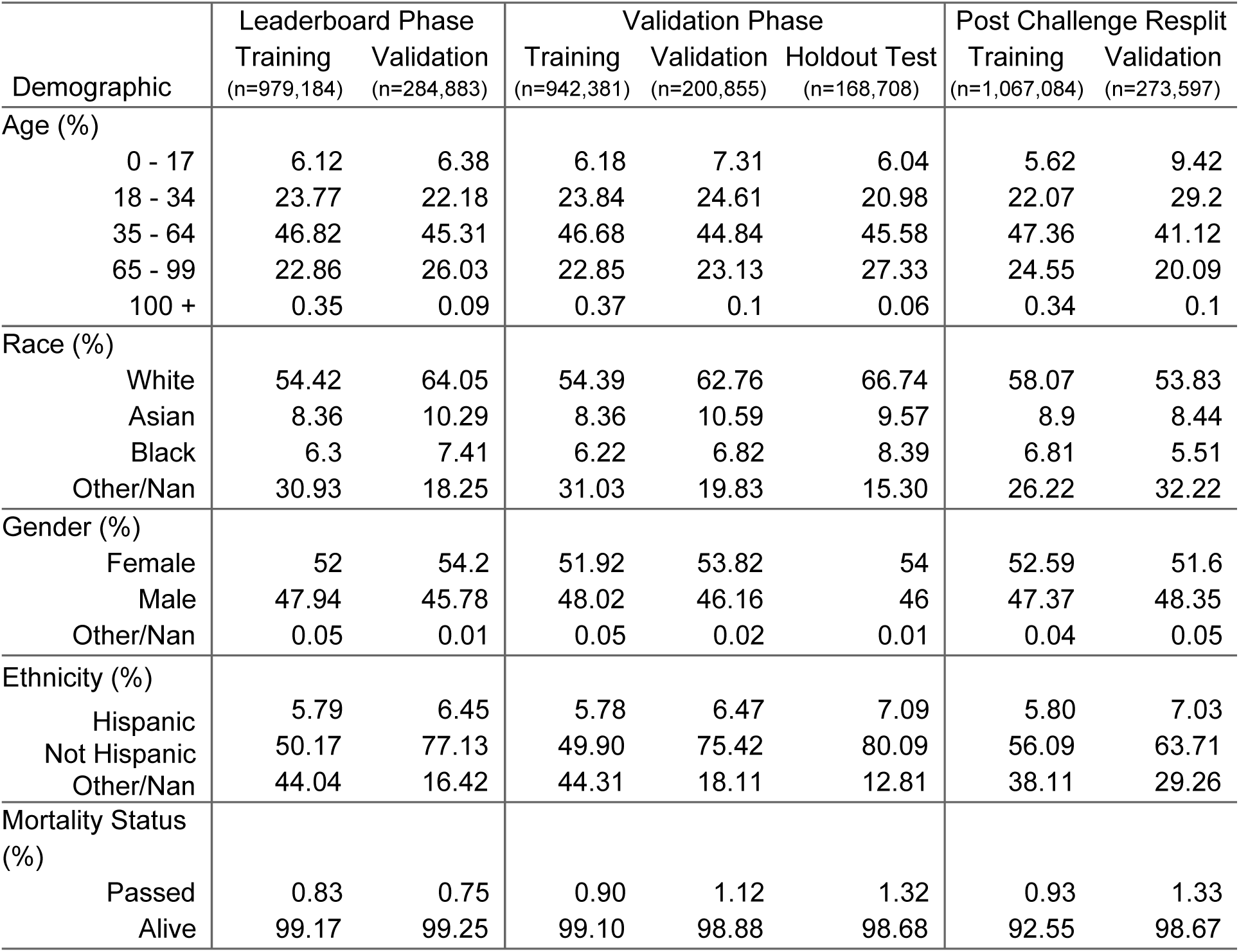
Demographic makeup as a percentage of the individual data sizes across the different versions of data used in the DREAM Challenge. All values represent the percentage of the total number of patients in the dataset of interest. We include a 100+ category as a standalone age category because that age range is of questionable quality. This gives some idea to the quality of the data made available.

### Model evaluation

#### Challenge Evaluation Metrics

The Area Under the Receiver Operator Curve (AUROC) was used as the primary metric for assessing model performance. A Bayes Factor, *K*, (bootstrapped distributions *n* = 10,000) was computed to determine if the AUROCs between two models were consistently different. If two models were found to have a small Bayes Factor (*K < 19*), we used the Area Under the Precision Recall Curve (AUPRC) as a tie-breaking metric. Both the AUROC and AUPRC were computed for all submissions and were used to rank teams on the Challenge leaderboard.

During the leaderboard phase, models were scored against the leaderboard validation data to build the initial leaderboard phase model ranking. During the final validation phase, models were scored against the prospectively-collected holdout test set. The top performing teams of the EHR DREAM Challenge: Patient Mortality were declared from the resulting validation phase model rankings. This holdout test set served to evaluate models on prospectively collected clinical data, testing a models ability to generalize over shifting clinical practice and data collection.

#### Re-split Data Validation

During the challenge, we used the validation phase holdout test set to build a final ranking of model performance. This holdout set only contained patients who appeared in the UW medical system in 2019. This left a 6-month longitudinal gap between the end date of the training data and the start date of the holdout test data. We combined all the datasets (training, validation, and holdout test) and re-divided the dataset into an 80/20 split between training and testing data using the same prospective splitting method we used to split the initial leaderboard data (details in the supplemental material). We trained the models on the 80% training data and evaluated the trained models against the 20% test data. This allowed us to compare the effect of the 6-month gap on model performance.

#### Subpopulation Accuracy Comparison

We evaluated model performance across various subpopulations which were defined by different demographic or clinical features including race, gender, ethnicity, age, and type of last visit. We compared model accuracy for each subpopulation against the accuracy of every other subpopulation within the same demographic or clinical group, calculating a paired Bayes Factor to evaluate the magnitude of accuracy differences. We ran this experiment on the prospectively gathered validation phase data.

#### Model Features

The top 5 performing teams were asked to output a list of features with accompanying weights in order to assess which features were most important in their models. We gathered a list of information that included the name of the feature, the “concept_id” s that were used to build those features, along with a weight or score that indicated the impact of that feature.

## Results

The EHR DREAM Challenge on all-cause patient mortality prediction was held between September 9, 2019 and February 23, 2020. Participants were asked to submit software programs that the Challenge organizers - acting as an honest broker - applied to hidden EHR data for training and model validation (Figure 1, Supplemental Figure 2). Data was split into training, validation, and holdout testing data sets that were used across two phases of the Challenge: a leaderboard phase and a final validation phase. Within the final validation phase, 942,381 patients were available for model training and 168,708 patients were used for model validation, with mortality rates of 0.90% and 1.32% respectively (Table 1). The Challenge received a total of 132 submissions from 25 teams that were able to be successfully executed with valid predictions.

During the leaderboard phase, of the 25 successfully validated models, ten teams exceeded AUROC > 0.9. AI4Life led the leaderboard phase - achieving an AUROC = 0.979 (0.977, 0.981) and AUPR = 0.614 (Table 2). In the final validation phase, 15 teams submitted successfully validated models, with three teams achieving an AUROC > 0.9 (Table 2, Figure 2). The top performing team, UW-biostat, achieved an AUROC=0.947 (0.924,0.952) and an AUPR=0.478. Between the leaderboard and validation phases, the average decrease in AUROC was 0.069 with the top 5 models decreasing by an average of 0.024 and the bottom 8 models decreasing by an average of 0.10. The top 5 models from the validation phase were ranked second, third, 11th, 10th, and 7th respectively at the end of the leaderboard phase but were ranked in the top 5 due to having the lowest decrease in performance.

**Figure 2.**
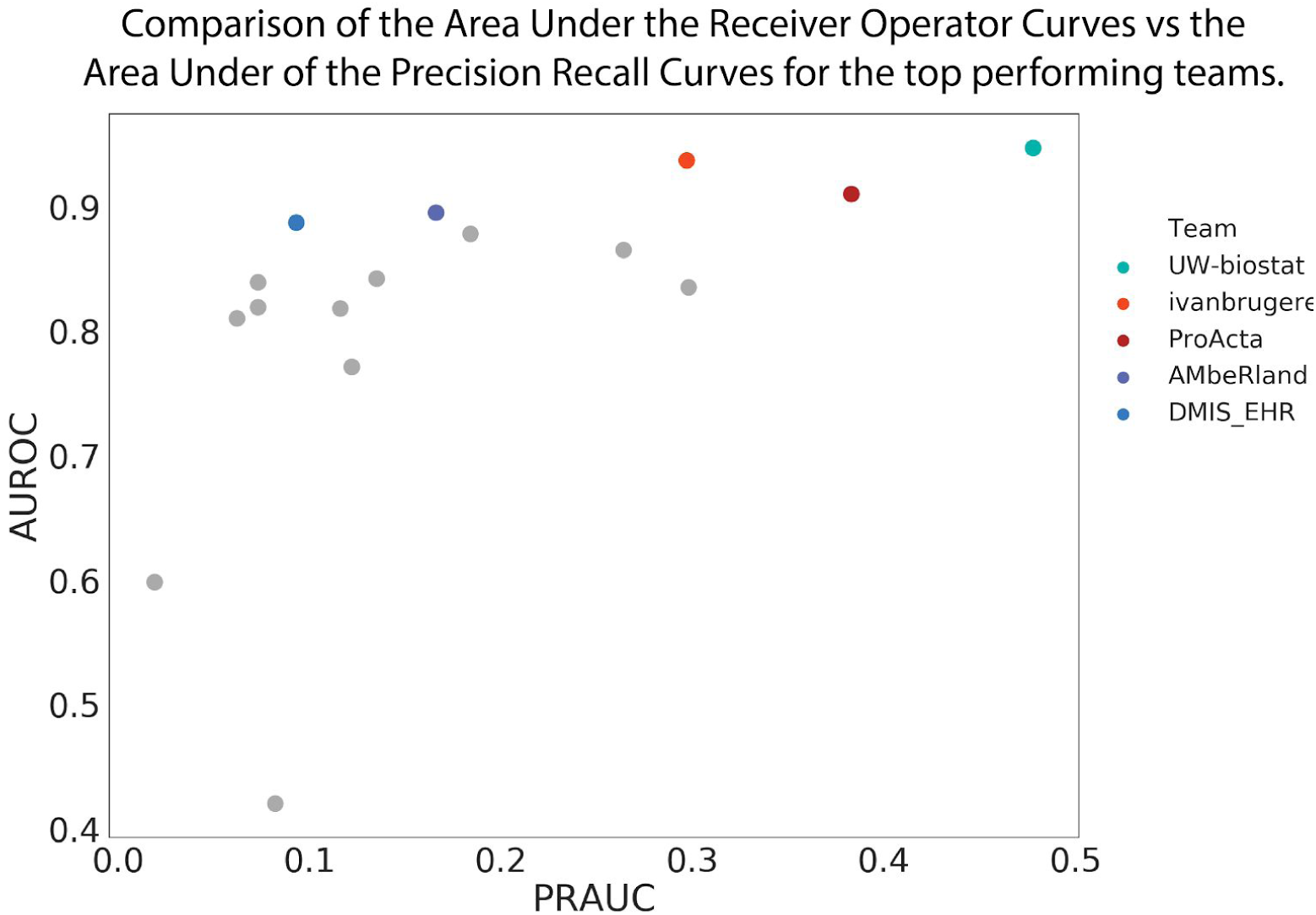
The Receiver Operator Curves and the Precision Recall Curves from all the models submitted in the validation phase. The top 5 models were not the top 5 models in the evaluation phase, but were more robust to longitudinal changes in the data. AUROCs and AUPRCs are reported in Table 1 for the leaderboard data, validation data, and the Resplit data. Comparison of leaderboard phase to validation phase scores are found in Figure 5.

**Table 2.** Top 15 teams and the metrics for their highest performing models. 95% confidence intervals were calculated using bootstrapped (n=1000) distributions. The Delong test p value was generated by comparing each team’s model with the team’s model ranked below them. Leaderboard phase scores were generated using the models submitted during the final validation phase.

| Team | Leaderboard Phase |  |  | Validation Phase |  |  |  | Post Challenge Resplit |  |  |
| --- | --- | --- | --- | --- | --- | --- | --- | --- | --- | --- |
|  | AUROC | AUROC 95% CI | AUPR | AUROC | AUROC 95% CI | Delong p value | AUPR | AUROC | AUROC 95% CI | AUPR |
| UW-biostat | 0.972 | (0.969, 0.975) | 0.524 | 0.947 | (0.942, 0.951) | 1.70E-04 | 0.478 | 0.964 | (0.961, 0.967) | 0.43 |
| Ivanbrugere | 0.968 | (0.964, 0.971) | 0.474 | 0.938 | (0.933, 0.942) | 1.96E-07 | 0.3 | 0.956 | (0.953, 0.96) | 0.409 |
| ProActa | 0.943 | (0.937, 0.948) | 0.458 | 0.91 | (0.903, 0.918) | 2.84E-03 | 0.383 | 0.904 | (0.898, 0.91) | 0.43 |
| AMbeRland | 0.942 | (0.937, 0.947) | 0.288 | 0.897 | (0.89, 0.903) | 4.18E-02 | 0.163 | 0.929 | (0.924, 0.934) | 0.284 |
| DMIS_EHR | 0.915 | (0.91, 0.92) | 0.111 | 0.887 | (0.88, 0.89) | 5.95E-02 | 0.093 | 0.939 | (0.936, 0.943) | 0.347 |
| PnP_India | 0.958 | (0.954, 0.963) | 0.449 | 0.876 | (0.87, 0.883) | 1.26E-01 | 0.182 | - | - | - |
| ultramangod671 | 0.882 | (0.874, 0.891) | 0.289 | 0.865 | (0.856, 0.875) | 2.45E-03 | 0.264 | 0.868 | (0.86, 0.876) | 0.37 |
| HELM | 0.951 | (0.948, 0.955) | 0.323 | 0.842 | (0.834, 0.85) | 5.65E-01 | 0.135 | - | - | - |
| AI4Life | 0.979 | (0.977, 0.981) | 0.614 | 0.831 | (0.82, 0.841) | 5.28E-01 | 0.302 | 0.971 | (0.969, 0.974) | 0.63 |
| Georgetown - ESAC | 0.938 | (0.933, 0.942) | 0.168 | 0.839 | (0.832, 0.848) | 1.64E-02 | 0.073 | 0.938 | (0.933, 0.941) | 0.272 |
| LCSB_LUX | 0.956 | (0.952, 0.959) | 0.307 | 0.82 | (0.81, 0.829) | 8.41E-01 | 0.116 | 0.936 | (0.932, 0.94) | 0.201 |
| QiaoHezhe | 0.925 | (0.92, 0.93) | 0.16 | 0.819 | (0.81, 0.827) | 2.92E-01 | 0.073 | - | - | - |
| chk | 0.903 | (0.896, 0.908) | 0.159 | 0.808 | (0.8, 0.817) | 1.38E-05 | 0.062 | 0.811 | (0.804, 0.818) | 0.061 |
| moore | 0.955 | (0.951, 0.958) | 0.313 | 0.771 | (0.757, 0.784) | 9.51E-45 | 0.122 | 0.947 | (0.943, 0.95) | 0.377 |
| tgaudelet | 0.904 | (0.898, 0.91) | 0.278 | 0.807 | (0.798, 0.817) |  | 0.201 | 0.158 | (0.151, 0.166) | 0.007 |

Teams used a variety of machine learning techniques in their submitted models. Of the 15 validated models, 12 were boosted methods (LightGBM ^23^, XGBoost ^24^, CatBoost ^25^, Generalized Boosted Regression ^26^), 2 were logistic regression, and 1 was a neural network (Supplemental Table 3). Of the top 5 models, 2 were LightGBM, 1 was logistic regression, 1 was CatBoost, and 1 was Generalized Boosted Regression. Each model used a different feature selection method ranging from randomly sampling all available concepts (Team IvanBrugere), carefully selecting a few features from the literature (Team LCSB_LUX), and using the structure of the concept ontologies to roll up low-level granular concepts into broad categories of disease and drugs as features (Team UW-biostat). We developed an ensemble model with the top performing models but observed that the ensemble did not meaningfully improve prediction accuracy over the top models (see Supplementary materials).

### Top performing model

UW-biostat’s (University of Wisconsin-Madison, Biostatistics and Medical Informatics) model achieved the highest Area Under the Recall curve during the final validation phase. While they were not the highest scoring model during the leaderboard phase, their model had the smallest decrease in performance of any model between the leaderboard phase and the validation phase, leading to their model having the highest score during the validation phase. The team used ontology-rollup to reduce feature dimensionality and used time binning and sample reweighting to capture longitudinal characteristics. For model development, they trained and tuned a LightGBM model to predict the mortality risk of each patient. To take into account potential data drift in EHRs ^4,27–29^, the team upweighted more recent patients during optimization and training of their model. In order to validate the model’s “future-proof” ability, they ordered the labeled patients by their last visit date from recent to early and used the top 15% of patients for validation. A more detailed description of this model and of the top 5 models are provided in the supplementary material. Model code can be found at https://github.com/GGGGFan/A-LightGBM-Model-to-Predict-180-Days-Mortality-Using-EHR-Data.

### Demographic Evaluation

To assess whether models generalized across patient subpopulations, we evaluated model accuracy across multiple demographic and clinical groups including race, gender, age, ethnicity, and last visit type. For each demographic or clinical strata, we generated bootstrapped distributions (*n* = 10,000) for each demographic strata in each model and ran paired permutation tests, calculating Bayes factors to assess the level of evidence for performance differences between subpopulations.

Models were consistently more accurate on Asian patients when compared to any other racial group (Figure 3, Supplemental Table 2), despite Asian patients only making up 8.4 percent of the validation data and 9.6 percent of the validation phase training data (Table 2). Methods varied in their accuracies for other races with some models (*e*.*g*. UW-biostat, IvanBrugere, Proacta, AMbeRland, DMIS_EHR) scoring higher on White patients compared to Black patients, and others scoring higher on Black patients than White patients (PnP_India, HELM, Georgetown-ESAC, AI4Life) (Figure 3, Supplemental Table 2).

**Figure 3.**
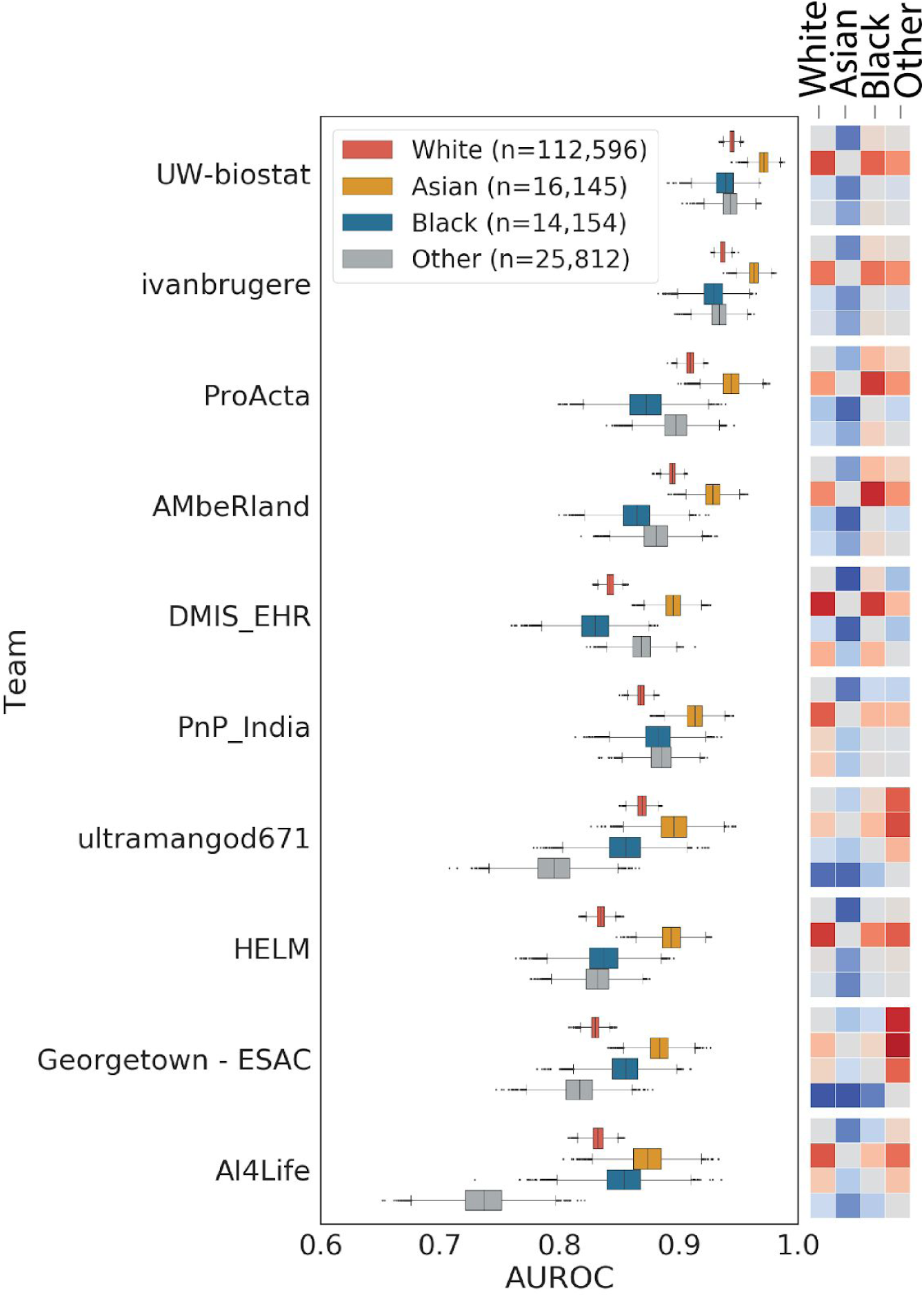
Bootstrapped distributions (*n* = 10,000) of the top 10 model AUROCs broken down by race. Model predictions were randomly sampled with replacement and scored against the benchmark gold standard. Comparisons were made between each category of race and Bayes values calculated to assess the level of evidence for the model having a higher accuracy on racial category compared to another category. The heat maps represent the log of the calculated Bayes factors when comparing racial groups within each model. The darker the red, the stronger the evidence for the racial category being higher than the comparison category. Bayes factor values range from 10000 to 0.0001. The darker the blue, the stronger the evidence for the racial category being lower than the comparison category. The color scale is normalized across all comparisons. Raw Bayes factor values can be found in Supplemental Table 2.

Without exception, models were more accurate on female patients than on male patients with Bayes factors greater than ten (strong evidence) for nine of the top 15 models (Supplemental Figure 4). As the challenge asked participants to predict mortality status 180 days from the last visit, we examined whether there were differences in model performances based on whether the last visit was inpatient, outpatient, or an emergency room visit. Most models had lower accuracy when the last visit was an outpatient visit, with the exception of 3 models (ultramangod671, Georgetown - ESAC, AI4Life in Figure 4). On patients where the last visit was an emergency room visit, models showed a wide variety of accuracies. In a few cases, models that had an overall lower model accuracy had higher accuracies on patients in the emergency room (compare ProActa to PnP_India in Figure 4).

**Figure 4.**
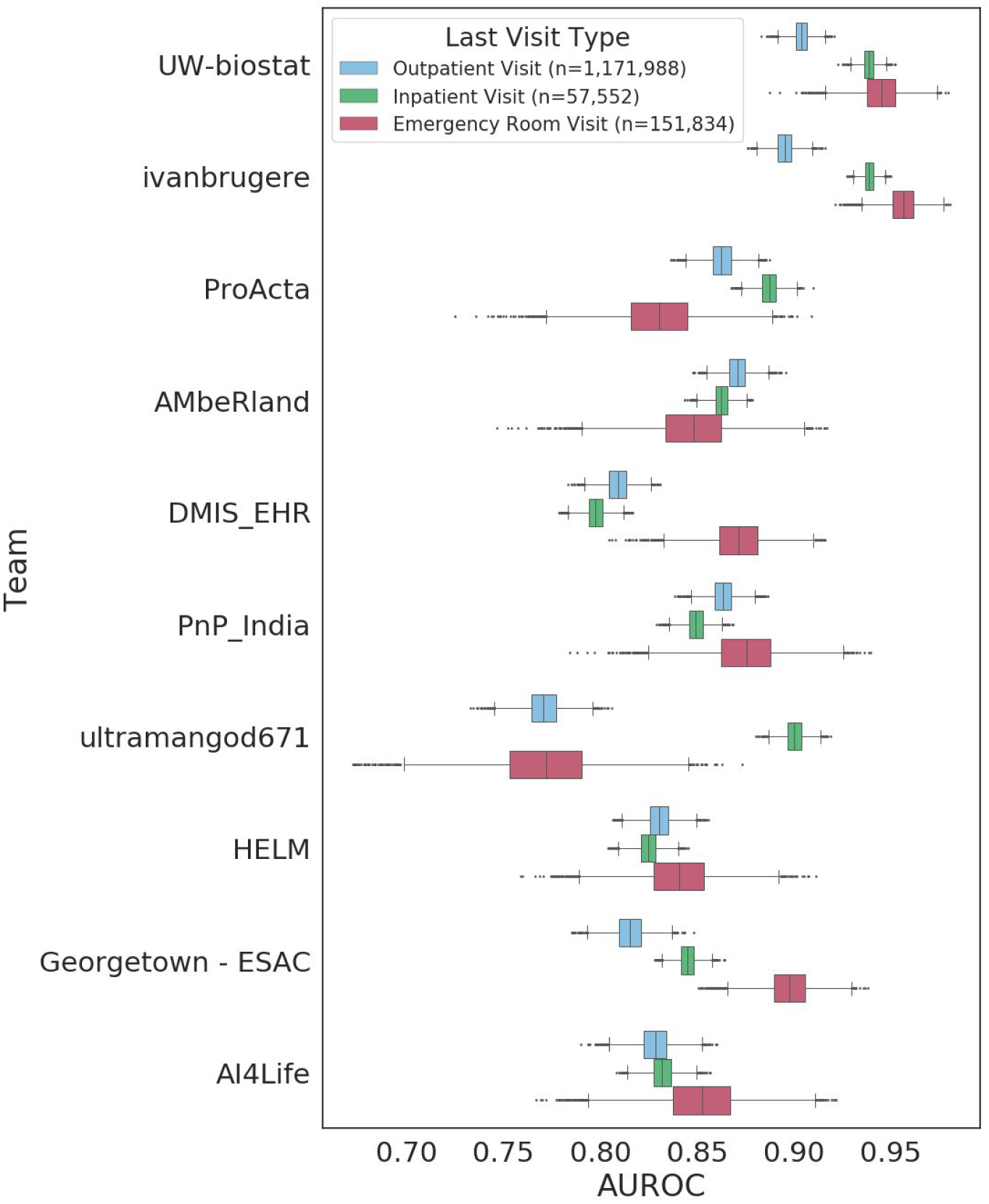
Bootstrapped distributions (n=10,000) of the top 10 model AUROCs broken down by the last visit type of the patients. The visit types evaluated were Inpatient Visits, Outpatient Visits, Emergency Room visits.

### Assessment of important features

The top five highest scoring teams were asked to adjust their dockerized models to output their trained features as a list of codes/values with associated weights from their trained models. In order to compare features across models, these teams reported which terms (SNOMED, RxNorm, LOINC, etc.) were used during any feature engineering and submitted brief descriptions of those features. The feature count and number of concepts used are reported in the supplemental materials (Supplemental Table 3).

The top four teams were able to successfully extract the features and weights from their models and output them into a human readable file. Table 3 reports the top 10 features of each model, including engineered features (i.e. presence or absence of a category of diagnosis or drug) and raw concepts from the data (i.e. granular SNOMED or LOINC codes). Some of the highly weighted features included the age of the patient at their last visit, systolic and diastolic blood pressure, heart rate, and a code for Do Not Resuscitate.

**Table 3.** The top 10 weighted features as reported from the top 4 performing teams. Features were ordered by their model weight and assigned a rank out of all available features. Feature names were either reported by the teams or were mapped using the OMOP concept table from the reported concept ids.

| Rank | UW-biostat | IvanBrugere | ProActa | AMbeRand |
| --- | --- | --- | --- | --- |
| 1 | Age | Not for resuscitation | Age | Age |
| 2 | Average pulse | Temperature | Creatinine in Serum | Not for resuscitation |
| 3 | Average Diastolic Blood Pressure | Albumin in Plasma | Heart rate | Antineoplastic chemotherapy regimen |
| 4 | Average Systolic Blood Pressure | History of clinical finding in subject | Inpatient Visit | Lactate dehydrogenase (LD), (LDH) |
| 5 | Latest Systolic Blood Pressure value | Natriuretic peptide B [Mass/volume] in Serum or Plasma | Blood typing, serologic; ABO | Administration of antineoplastic agent |
| 6 | Year of last visit | Racial Variable (White) | Palliative care | Patient encounter procedure |
| 7 | Latest pulse measurement | Antineoplastic chemotherapy regimen | Albumin in Plasma | Secondary malignant neoplasm of lung |
| 8 | Latest Diastolic Blood Pressure Value | Protein in Plasma | Hematocrit of Blood by Automated count | Disorder of lung |
| 9 | Latest Glucose measurement | Heart rate | Neutrophils/100 leukocytes in Blood by Automated count | Dexamethasone |
| 10 | Indicator: Unknown conditions | Essential hypertension | Cholecalciferol | Bacterial culture |

## Discussion

Machine learning models are increasingly regarded as foundational to any precision medicine strategy. The assessment of model accuracy and utility in a healthcare environment is challenged by limited data availability, concerns about breach of protected health information confidentiality, and lack of technical infrastructure, domain expertise and process to systematically manage model evaluations. We implemented an architecture for an unbiased and transparent assessment of methods that overcomes these limitations, and in doing so were able to improve existing methodology. We demonstrated how community challenges can provide an inclusive and rigorous environment for hosting a machine learning clinical trial.

Using the Model-to-Data framework, 25 international teams submitted machine learning models to a private clinical dataset that otherwise would have remained inaccessible to these researchers. This was enabled by leveraging a common data model, in this case OMOP, a synthetic dataset for technical development and validation, a cloud environment hosting the synthetic data for pipeline and execution evaluation, standard containerization software, and a secure environment hosting the private clinical data.

For the first EHR DREAM Challenge, we asked participants to predict 180-day mortality status of patients in the University of Washington clinical records. However, this prediction question is not immune to censoring, and is susceptible to an open world limitation as some patients may die out of state or outside UW clinical care without the ability to map their death to UW clinical records ^30^. Another limitation, as shown both in the selected features (Table 3) and in model accuracy across last visit type (Figure 4), is that all-cause mortality is not a clinically actionable question, as models trained for all-cause mortality are not specific enough for clinical action.

Given that some of the highlighted features used to predict death included age, palliative care, designation for do not resuscitate, and treatments or diagnoses of cancer (neoplasms), many of the models developed in this challenge were identifying the most obvious patients in the UW population. However, the highest performing model assigned the most weight on specific measurements and their values, not just the presence or absence of a measurement or condition, and was utilizing the hierarchical design of the available biomedical ontologies to “roll up” low level granular codes. This points to the fact that the Model-to-Data framework can be used in concert with intelligent feature engineering and selection.

Assessing a wide variety of methods from teams allowed us to evaluate the best approaches and assess inter-method variability when holding the evaluation data constant. Interestingly, even though models were trained and evaluated on the same data, there was variance in model accuracy across different demographic groups. White, Black, and other racial groups showed differences across models, with some models scoring higher on Black patients than White patients and vice versa, while Asian patients were consistently more accurate across nearly all models. This may have to do with the cause of death, as prevalence of different causes of death may vary between different populations. Unfortunately, we did not have access to cause of death data at the time of this analysis. With the exception of the 0-17 age group, method accuracy was inversely correlated with age. One hypothesis for this trend is that younger patients who pass away in 180 days and are coming into the hospital are more likely to have extreme conditions and have a higher risk of death, while older patients are simply more likely to have diseases and health problems in general, making it more difficult to predict risk of death.

Models also had varied accuracies from the last visit type, highlighting the need to develop context-specific clinical prediction algorithms, although the two most accurate models were still more accurate on outpatient visits than all other less accurate models (Figure 4). In other cases, models were aligned in their bias, universally scoring higher on females than males (Supplemental Figure 4). We found no meaningful difference in model accuracy between Hispanic and Non-Hispanic ethnicity. (Supplemental Figure 4).

Evaluating models in a pseudo-prospective manner allowed us to assess how models would perform over time in the UW environment. We found that most models decreased in performance in the validation phase when compared to the leaderboard phase (Figure 5). This is in line with the literature, as previous studies have shown that the utility of clinical data can have a half-life of as little as three months ^4^. As an example from the UW data, 19.5% of patients with a condition record in 2018 had the concept code for “malignant tumor of prostate” while in 2019, only 1.5% of patients with a condition record had that code. Comparing results from the post-challenge resplit data to the validation phase final results, the majority of models performed better when the training data was longitudinally closer to the test data (Table 2).

**Figure 5.**
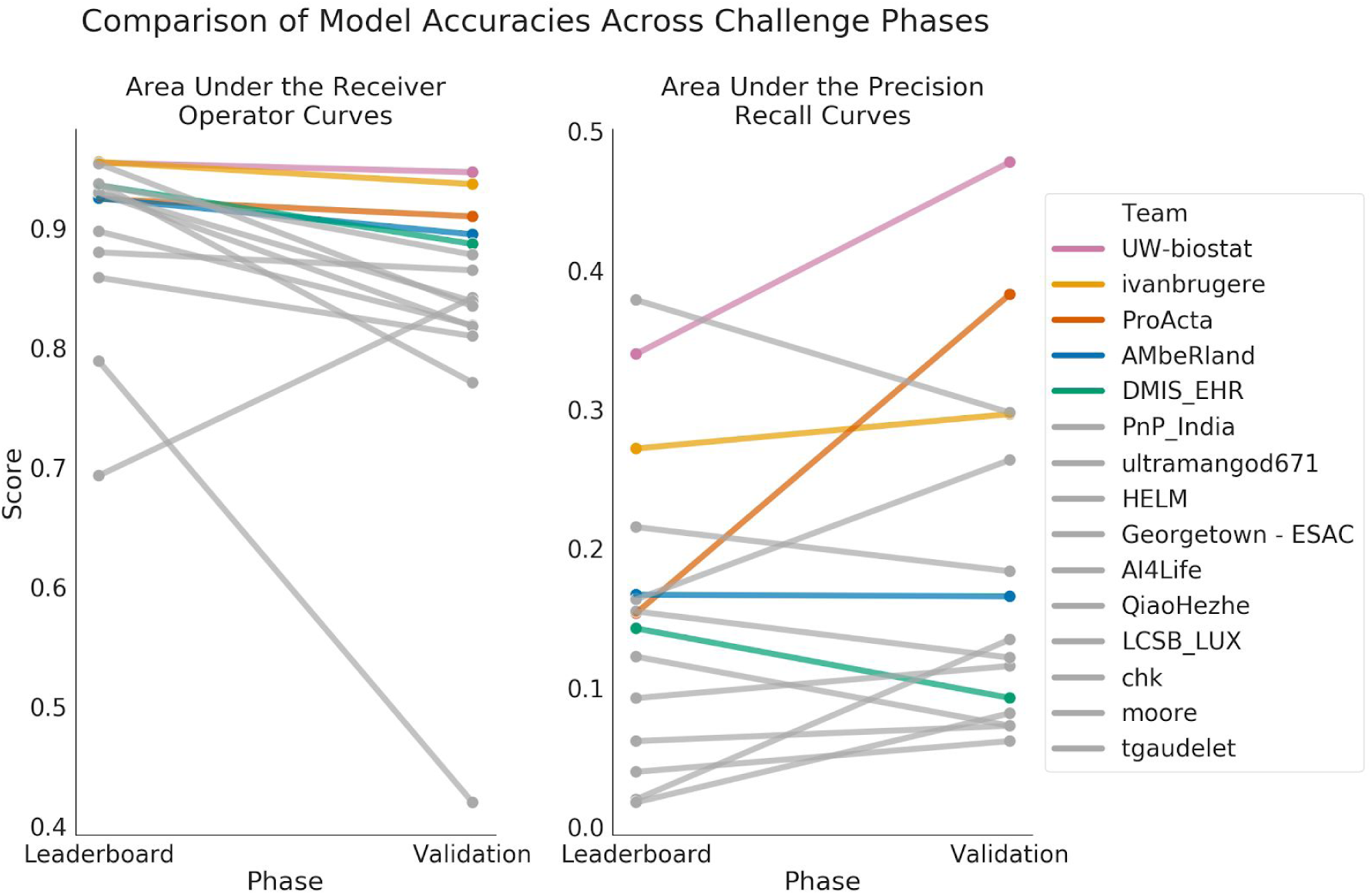
Comparison of model performance between the leaderboard phase and the validation phase. All models decreased in AUROC except for HELM while many models increased in AUPRC. The top 5 team AUROCs decreased the least between the two phases.

UW-biostat explicitly down-weighted older data and relied more heavily on the most recent 6-months of data in the training dataset resulting in their model having the lowest decrease in performance of any model. By combining this technique with their “roll up” feature engineering, UW-biostat’s model proved to be the most robust against longitudinal changes and covariate shift. In contrast, some models dropped by a significant margin. For instance, AI4Life’s model was among the most accurate during the leaderboard phase, but dropped to tenth in the validation phase (Table 2). While it is difficult to completely account for their drop, one possible explanation is their overall lower accuracy on first time visiting patients (Supplemental Figure 7) combined with the increase of first time visitors in the validation data (leaderboard phase data - 13.8% compared to the validation phase data - 19.7%). AI4Life’s model had a high score on the resplit data, indicating that their model was susceptible to covariate drift as well. Evaluating models prospectively or pseudo-prospectively evaluates models in the manner in which they will be used and brings us closer to understanding how the models will perform in a live clinical setting.

### Limitations

For this challenge, we set a submission limit of 10 hours for both the training and testing stages of model runtime. While this was implemented to limit the burden to the University of Washington secure servers, this also limited the types of models participants were able to build and excluded deep learning or more sophisticated, time consuming models. However, limiting the time also forced participants to carefully consider the efficiency of their algorithms so extraneous calculations or operations were not run.

Training and evaluating on data from a single site limited our ability to control for overfitting. While we did prospectively evaluate models on a future holdout set to try and control for overfitting, evaluation on data from one site does not properly assess model generalizability. For future assessments, we hope to partner with other hospitals to externally validate models.

## Conclusion

Machine learning promises to enhance patient care and improve health outcomes; however, if not properly vetted and evaluated, risks and negative effects may be introduced. These risks include breach of privacy in the development and assessment of methods, inaccuracy or methodological bias when deployed, and the gradual loss of accuracy over time as data and business practices change. This study highlights these challenges by showing that while highly accurate methods are possible, even methods from world-class scientists have considerable variability, and that variability (such as differences in accuracy based on race or gender) may not be detectable from high-level measures such as population-level AUCs. Our framework enables this assessment and also brings the community challenge culture to private datasets, in this case data that is subject to the HIPAA privacy rule. Further, machine learning methods may be able to address some causes of treatment disparities but may cause others for patients without rich longitudinal data or patients of certain races, gender, or age. Based on these results, we believe that multisite standardized architecture and independent oversight is required to truly assess new methods.

## Data Availability

Due to privacy concerns, the clinical data used in this study can not be made available.

## Acknowledgements

This work was supported by the Clinical and Translational Science Awards Program National Center for Data to Health funding by the National Center for Advancing Translational Sciences at the National Institutes of Health (grant numbers U24TR002306 and UL1 TR002319). Any opinions expressed in this document are those of the Center for Data to Health community and the Institute for Translational Health Sciences and do not necessarily reflect the views of the National Center for Advancing Translational Sciences, team members, or affiliated organizations and institutions. TB, YY, SM, JG, TY, TS, and JP were supported by grant number U24TR002306. TB, JP, and SM were supported by grant number UL1 TR002319.

## Patient Mortality Prediction DREAM Challenge Consortium Authors

Aaron Lee, MD, MSc (Department of Ophthalmology, University of Washington); Ali Salehzadeh-Yazdi, PhD (Department of Systems Biology and Bioinformatics, University of Rostock, Rostock, Germany); Anand Basu (ESAC Inc.); Anas Belouali, MEng, MS (Innovation Center for Biomedical Informatics (ICBI), Georgetown University, Washington DC); Ann-Kristin Becker (Institute of Bioinformatics, University Medicine Greifswald); Ariel Israel, MD PhD (Department of research and data, Division of Planning and Strategy, Clalit Health Services, Tel-Aviv, Israel); B. Winter, ツ (ICM, Universitätymedizin, Greifswald, Mecklenburg-Vorpommern, Germany); Carlos Vega Moreno, PhD (Bioinformatics Core, Luxembourg Centre for Systems Biomedicine (LCSB), University of Luxembourg, Esch-sur-Alzette, Luxembourg); Christoph Kurz, PhD (Helmholtz Zentrum München, Institute of Health Economics and Health Care Management, Neuherberg, Germany; Munich School of Management and Munich Center of Health Sciences, Ludwig-Maximilians-Universität München, Munich, Germany); Dagmar Waltemath, PhD (Medical Informatics, Institute for Community Medicine, Greifswald, Germany, MV); Darius Schweinoch (Institute of Bioinformatics, University Medicine Greifswald, Greifswald, Mecklenburg-Vorpommern, Germany); Enrico Glaab, PhD (University of Luxembourg, Luxembourg Centre for Systems Biomedicine, Esch-sur-Alzette, Luxembourg); Gang Luo, PhD (Department of Biomedical Informatics and Medical Education, University of Washington); Helena U. Zacharias, PhD (Department of Psychiatry and Psychotherapy, University Medicine Greifswald, Greifswald, Germany); Hezhe Qiao, PhD (Chongqing Institute of Green and Intelligent Technology, Chinese Academy of Sciences, Chongqing, China.); Julia Truthmann, PhD (Department of Family Medicine, Institute for Community Medicine, University Medicine Greifswald); Kari A. Stephens, PhD (Department of Psychiatry and Behavioral Sciences, University of Washington); Lars Kaderali, Dr. (Institute of Bioinformatics, University Medicine Greifswald, Greifswald, MV, Germany); Lav R. Varshney, Ph.D. (Salesforce Research, Palo Alto, CA, USA; University of Illinois at Urbana-Champaign, Urbana, IL, USA); Marcus Vollmer, Dr. rer. nat. (Institute of Bioinformatics, University Medicine Greifswald, Greifswald, Germany; DZHK (German Centre for Cardiovascular Research), partner site Greifswald, Germany); Maria-Theodora Pandi (Laboratory of Pharmacogenomics and Individualized Therapy, Department of Pharmacy, School of Health Sciences, University of Patras, Rion, Greece); Martin L. Gunn, M.B.Ch.B. (Department of Radiology, University of Washington); Meliha Yetisgen, PhD (Department of Biomedical Informatics and Medical Education, University of Washington); Michael Holck (ESAC, Inc.); Neetika Nath, PhD (University medicine Greifswald Department of Bioinformatics 17475 Greifswald); Noah Hammarlund, PhD (Department of Biomedical Informatics and Medical Education, University of Washington); Oliver Müller-Stricker (Institute of Bioinformatics, University Medicine Greifswald, Greifswald, Germany); Panagiotis Togias (Laboratory of Medical Physics, School of Health Sciences, Department of Medicine, University of Patras, Rion, Greece); Patrick J. Heagerty, PhD (Department of Biostatistics, University of Washington); Peter Banda, PhD (Bioinformatics Core, Luxembourg Centre for Systems Biomedicine, University of Luxembourg, Esch-sur-Alzette, Luxembourg); Peter Muir, MD (PJM Consulting LLC, ESAC Team); Roland Krause, Dr. sc. hum. (Bioinformatics Core, Luxembourg Centre for Systems Biomedicine (LCSB), University of Luxembourg, Esch-sur-Alzette, Luxembourg); Ron Henkel (Medical Informatics Section Epidemiology of Health Care and Community Health University Medicine Greifswald); Sagar Madgi (Associate Principal, Advanced Data Science Track, ZS Associates, Bengaluru, Karnataka, India); Samir Gupta (Innovation Center for Biomedical Informatics, Georgetown University, Washington, DC, USA); Shabeeb Kannattikuni (Innovation Center for Biomedical Informatics, Georgetown University Medical Center, Washington DC, USA); Shamim Sarhadi, MSc (Department of Medical Biotechnology; Faculty of Advanced Medical Sciences; Tabriz University of Medical Sciences; Tabriz; Iran.); Shikhar Omar (Data Science Team, ZS Associates, Bengaluru, Karnataka, India); Shuo Wang (Georgetown University); Soumyabrata Ghosh, PhD (Bioinformatics Core, Luxembourg Centre for Systems Biomedicine (LCSB), University of Luxembourg, Esch-sur-Alzette, Luxembourg); Stefan Simm, Assistant Professor (Institute of Bioinformatics, University Medicine Greifswald, Greifswald, Germany); Stefan Neumann (University Medicine Greifswald, Department of Bioinformatics, Greifswald, Mecklenburg-Vorpommern, Germany); Subha Madhavan, PhD, FACMI (Georgetown University); Thomas Von Yu, BA (Sage Bionetworks); Venkata Satagopam, PhD (Bioinformatics Core, Luxembourg Centre for Systems Biomedicine (LCSB), University of Luxembourg, Esch-sur-Alzette, Luxembourg); Vikas Pejaver, PhD (Department of Biomedical Informatics and Medical Education, University of Washington); Yachee Gupta (Medical Informatics Laboratory, Institute for Community Medicine, Section Epidemiology of Health Care and Community Health, University Medicine Greifswald, Germany.)

