## Supplemental Materials for "Evaluation of crowdsourced mortality prediction models as a framework for assessing AI in medicine"

### Supplemental Material

#### Data Creation Process

For this challenge, we wanted to evaluate models in a prospective manner, evaluating their performance over time on a live clinical data warehouse. However, we were technically limited and decided to run a pseudo-prospective challenge where we split the data into the training and evaluation data in a longitudinal manner where the newer data was part of the evaluation data and the older data is part of the training data. The last death record in the available UW OMOP repository at the time of this study was in February 2019. Any record or measurement that was found after this date was excluded from the challenge dataset. We wanted to split the data into an 80/20 ratio for training/evaluation. In order to do this, we defined a time range “evaluation window” in which if a patient had a visit in that window, we included them in the evaluation dataset. We expanded this evaluation window backwards in time until the number of included patients was 20% of the total data. The final evaluation window was approximately 9 months in length. We considered the date 180 days prior to the end of data (August 2018) the end of the “evaluation window” and the beginning of the evaluation window to be 9 months prior to the evaluation window start (November 2017). Patients who had visits outside the window, but none within the window, were included in the training data. Visit records that fell after the evaluation window end were removed from the evaluation dataset (Supplemental Figure 1, patient 7) and from the training dataset for patients who did not have a confirmed death (Supplemental Figure 1, patient 3). We only defined the true positives for the evaluation dataset and created a gold standard of these patients’ mortality status based on their last visit date and the death table. However, we gave the model developer the flexibility to select prediction dates for patients in the training dataset and to create corresponding true positives and true negatives for training purposes.

During the validation phase, a third dataset was created from patients who had visited the University of Washington medical system from January 2019 to June 2019. Patients who had a visit record in that time frame were added to the validation dataset, including patients who were previously in the leaderboard datasets (Supplemental Figure 1, Validation).

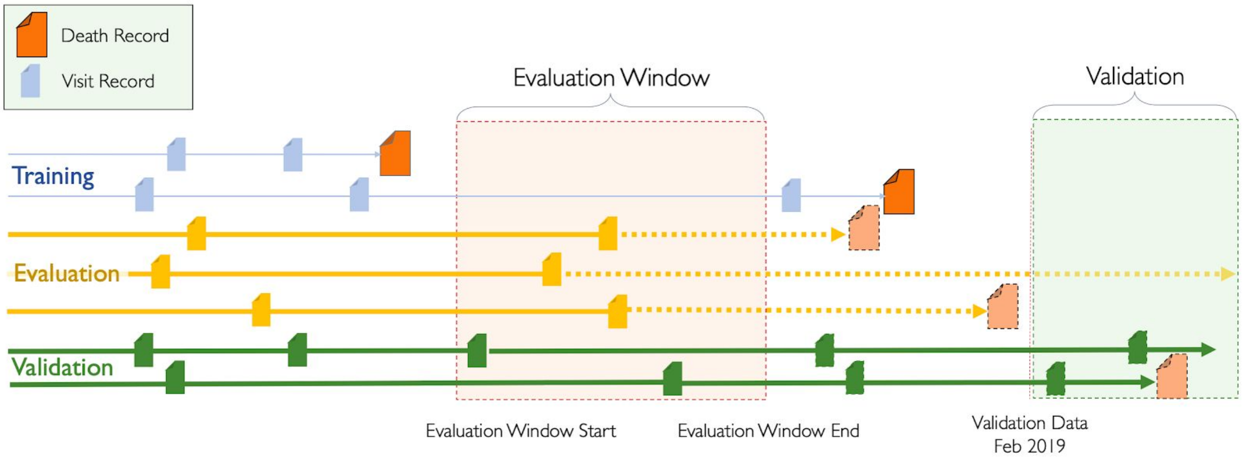


Supplemental Figure 1. Visualization of the data creation process. During the leaderboard phase, the available data was split into two datasets, training and evaluation, that was an 80/20 ratio. The evaluation data was defined by the evaluation window, which specified the time range in which patients who had a visit in that range were included in the evaluation dataset while all other patients were included in the training data. During the validation phase, patients who had visited the University of Washington medical system in 2019, after the end of the leaderboard data, were included in the validation data. This included patients who were previously in the leaderboard training and evaluation datasets.

### Synthetic Data

We derived a synthetic dataset from the SynPuf Synthetic OMOP dataset [20]. Starting with the original SynPuf dataset we adapted it to our challenge by randomly sampling terms that occurred more than 100 times from the University of Washington OMOP repository and then populating the tables of the original SynPuf dataset with these random samples such that the synthetic data closely resembled the UW OMOP data rather than the SynPuf data. This was to give participants an idea of what terms they could expect in the UW data. We also made a data dictionary available that included a list of these same codes.

During the original Model to Data pilot study[8], we found that external model developers had a hard time estimating their data curation and model runtimes using just the SynPuf dataset. This turned out to be caused by the difference in size and distribution of the synthetic data relative to the real UW data. We adjusted the size of the synthetic data by matching the distribution of records across real UW patients by first binning the UW patient population by the number of visit records they had in the UW data, then up sampling visit records of patients in the synthetic data until the number of patients in each of the bins matched the count in the UW data (i.e. upsampling synthetic visits until the number of patients with 100 visits matched the real UW data and the number of patients with 250 visits matched the real UW data, etc.).

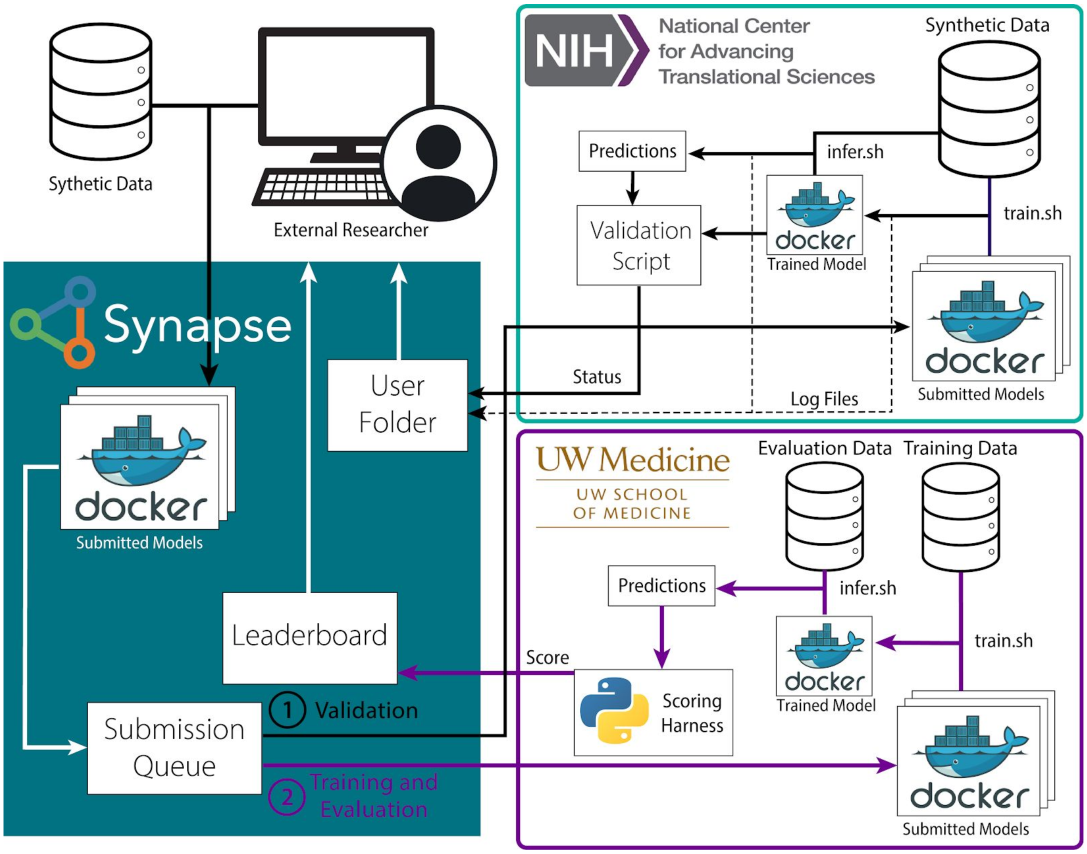


Supplemental Figure 2. The workflow of the challenge infrastructure. Participants developed a dockerized model using the synthetic data. These docker images were submitted to the Synapse collaboration platform to a submission queue. From this queue, images were pulled into a cloud environment hosting synthetic data where they were executed. Once the models were validated on synthetic data, they were pulled into the UW secure environment where they were executed and evaluated against the UW OMOP repository. Scores were returned to participants through the Synapse platform.

### Ensemble Model

To build an ensemble model, we train each participant's mortality prediction model on the validation phase training dataset and apply the trained models separately on the evaluation dataset and validation dataset to generate mortality probability for each patient in the corresponding dataset (evaluation dataset prediction and validation dataset prediction). An ensemble model (10-fold logistic regression model) takes as input evaluation dataset predictions from each model. The mortality outcome(deceased or not) for each patient in the evaluation dataset is used as the label. The ensemble model is validated using validation dataset predictions from individual models and gold standard for validation dataset. The ensemble model has a performance of ROC AUC: 0.888 and PR AUC: 0.372, which is worse than the best individual model(ROC AUC: 0.947, PR AUC: 0.478). This is because of the data difference between evaluation dataset and validation dataset when splitting in a prospective manner. Individual Model’s performance changes when performing on the evaluation dataset compared to validation dataset(see Supplemental Table 1). The ensemble model is trained on model’s prediction on evaluation dataset, however, model’s performance on evaluation dataset is not a good indicator for model’s performance on validation dataset.

Supplemental Table 1. Performance of trained models on each of the datasets used to develop the ensemble model.

| | **Evaluation** | | **Validation** | | **Rank** | |
|---|---|---|---|---|---|---|
| | ROC AUC | PRAUC | ROC AUC | PRAUC | Validation dataset | Evaluation dataset |
| UW-biostat | 0.972 | 0.524 | 0.947 | 0.478 | 1 | 2 |
| ivanbrugere | 0.968 | 0.473 | 0.937 | 0.297 | 2 | 3 |
| ProActa | 0.943 | 0.459 | 0.910 | 0.384 | 3 | 8 |
| AMbeRland | 0.942 | 0.288 | 0.895 | 0.166 | 4 | 9 |
| DMIS_EHR* | 0.915 | 0.111 | 0.887 | 0.093 | 5 | 12 |
| PnP_India | 0.958 | 0.449 | 0.878 | 0.184 | 6 | 4 |
| ultramangod671 | 0.882 | 0.288 | 0.865 | 0.270 | 7 | 15 |
| HELM | 0.951 | 0.306 | 0.842 | 0.126 | 8 | 7 |
| AI4Life | 0.979 | 0.614 | 0.835 | 0.298 | 9 | 1 |
| Georgetown-ESAC | 0.938 | 0.168 | 0.839 | 0.073 | 10 | 10 |
| LCSB_LUX | 0.956 | 0.308 | 0.818 | 0.116 | 11 | 5 |
| QiaoHezhe | 0.925 | 0.160 | 0.819 | 0.073 | 12 | 11 |
| chk | 0.903 | 0.159 | 0.810 | 0.063 | 13 | 14 |
| moore | 0.955 | 0.314 | 0.771 | 0.122 | 14 | 6 |
| tgaudelet | 0.904 | 0.129 | 0.420 | 0.030 | 15 | 13 |

### Challenge Timeline

The EHR DREAM Challenge had three phases: the open phase, the leaderboard phase, and the validation phase. We held the open phase from September 9, 2019 to October 9, 2019 and allowed participants to build and submit models to the cloud infrastructure hosting the synthetic data so challenge participants could become familiar with the process and the infrastructure. During the leaderboard phase, participants submitted models which were run on both the synthetic data and the UW EHR data. When models were pulled into the UW environment, they were first trained on the training dataset and tested against the evaluation dataset. The leaderboard phase was held from October 9, 2019 to January 28, 2020 in three rounds, each a month long. For each round, participants were allowed three valid submissions where a valid submission was a submission that passed the synthetic data validation and output a prediction file on the UW evaluation data that could be scored. Due to technical difficulties during the third round, we extended the third round 17 days, and gave participants two extra valid submissions. During the validation phase, participants submitted a final model that was first run on the synthetic data, then trained on the training dataset and tested on the validation dataset. The final scores were generated during the validation dataset and the top models were ranked based on their performance on the validation data. The validation phase was open for final submissions from February 4, 2020 to February 18, 2020. We announced the final results on May 4, 2020. The timeline with dates and labels is visualized in Supplemental Figure 3.

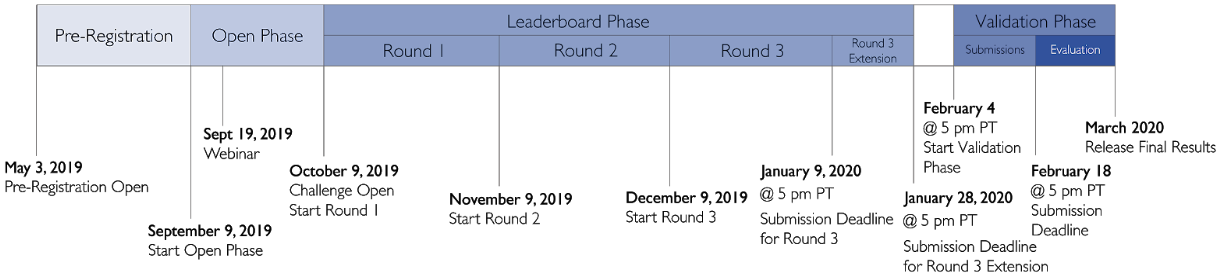


Supplemental Figure 3. Timeline of the EHR DREAM Challenge: Patient Mortality Prediction challenge. The challenge was held in three phases: open, leaderboard, and validation phase. The leaderboard phase was held in 3 rounds, each one month long, where each team was allowed up to three successful submissions.

Supplemental Table 2. Results of the permutation tests to evaluate the significance of the scoring distribution for each racial group from each submitted model. The values in each column represent the Bayes factors from the permutations. Each racial group was compared to each other with paired permutations (n=10,000) and the Bayes factor for each comparison were calculated.

| | | Paired Bayes Factor Comparisons | | | |
|---|---|---|---|---|---|
| Team | Race | White | Asian | Black | Other |
| | White | 0.94 | 1249.00 | 2.12 | 1.27 |
| | Asian | 1999.00 | 1.01 | 908.09 | 162.93 |
| | Black | 2.17 | 999.00 | 0.96 | 1.65 |
| UW-biostat | Other | 1.22 | 174.44 | 1.65 | 0.98 |
| | White | 0.98 | 587.24 | 2.71 | 1.53 |
| | Asian | 453.55 | 0.99 | 499.00 | 165.67 |
| | Black | 2.65 | 475.19 | 1.03 | 1.65 |
| ivanbrugere | Other | 1.65 | 165.67 | 1.66 | 1.01 |
| | White | 0.99 | 92.46 | 20.01 | 3.32 |
| | Asian | 99.00 | 0.97 | 3332.33 | 137.89 |
| | Black | 20.98 | 3332.33 | 1.02 | 4.86 |
| ProActa | Other | 3.35 | 125.58 | 4.73 | 0.99 |
| | White | 0.99 | 226.27 | 17.02 | 4.03 |
| | Asian | 195.08 | 0.98 | 4999.00 | 134.14 |
| | Black | 15.81 | 4999.00 | 1.03 | 2.77 |
| AMbeRland | Other | 4.05 | 152.85 | 2.73 | 1.00 |
| | White | 1.00 | 10000.00 | 2.89 | 30.25 |
| DMIS_EHR | Asian | 4999.00 | 1.03 | 3332.33 | 18.53 |

| | | | | | |
|---|---|---|---|---|---|
| | Black | 3.01 | 4999.00 | 1.00 | 22.42 |
| | Other | 32.78 | 18.65 | 23.51 | 0.98 |
| PnP_India | White | 0.98 | 1249.00 | 3.33 | 6.50 |
| | Asian | 1110.11 | 0.97 | 23.45 | 17.69 |
| | Black | 3.54 | 20.23 | 0.96 | 1.17 |
| | Other | 6.75 | 16.83 | 1.22 | 1.00 |
| ultramangod671 | White | 0.97 | 8.73 | 2.54 | 1110.11 |
| | Asian | 8.34 | 0.98 | 17.62 | 1999.00 |
| | Black | 2.72 | 16.92 | 0.99 | 24.64 |
| | Other | 2499.00 | 3332.33 | 24.97 | 0.99 |
| HELM | White | 0.99 | 4999.00 | 1.22 | 1.35 |
| | Asian | 3332.33 | 1.00 | 269.27 | 1110.11 |
| | Black | 1.22 | 293.12 | 1.01 | 1.43 |
| | Other | 1.33 | 768.23 | 1.39 | 1.00 |
| AI4Life | White | 1.00 | 20.88 | 3.62 | 4999.00 |
| | Asian | 20.14 | 1.02 | 3.25 | 10000.00 |
| | Black | 3.60 | 3.07 | 1.02 | 908.09 |
| | Other | 9999.00 | 10000.00 | 999.00 | 1.00 |
| Georgetown - ESAC | White | 0.96 | 908.09 | 7.41 | 3.03 |
| | Asian | 1427.57 | 0.95 | 13.93 | 587.24 |
| | Black | 7.50 | 12.48 | 0.99 | 11.02 |
| | Other | 3.07 | 433.78 | 11.30 | 1.04 |
| LCSB_LUX | White | 0.99 | 106.53 | 3.98 | 79.00 |
| | Asian | 112.64 | 0.99 | 7.73 | 9999.00 |
| | Black | 4.13 | 7.73 | 1.04 | 75.34 |
| | Other | 74.19 | 4999.00 | 64.79 | 0.99 |
| QiaoHezhe | White | 0.98 | 10000.00 | 3.74 | 10.31 |
| | Asian | 4999.00 | 1.03 | 80.97 | 38.84 |
| | Black | 3.77 | 73.07 | 1.01 | 1.34 |
| | Other | 9.57 | 37.61 | 1.43 | 1.00 |
| chk | White | 0.96 | 10000.00 | 2.11 | 1.20 |
| | Asian | 10000.00 | 0.98 | 3332.33 | 1110.11 |
| | Black | 2.12 | 9999.00 | 1.00 | 2.10 |
| | Other | 1.22 | 624.00 | 2.11 | 1.03 |
| moore | White | 1.00 | 13.93 | 9.65 | 10000.00 |
| | Asian | 13.10 | 0.98 | 1.09 | 10000.00 |
| | Black | 10.56 | 1.05 | 1.00 | 9999.00 |
| | Other | 10000.00 | 10000.00 | 10000.00 | 0.97 |
| tgaudelet | White | 0.96 | 1427.57 | 3.06 | 55.18 |
| | Asian | 1249.00 | 1.02 | 53.95 | 4999.00 |
| | Black | 3.18 | 48.26 | 0.98 | 40.67 |
| | Other | 58.17 | 10000.00 | 37.91 | 0.97 |

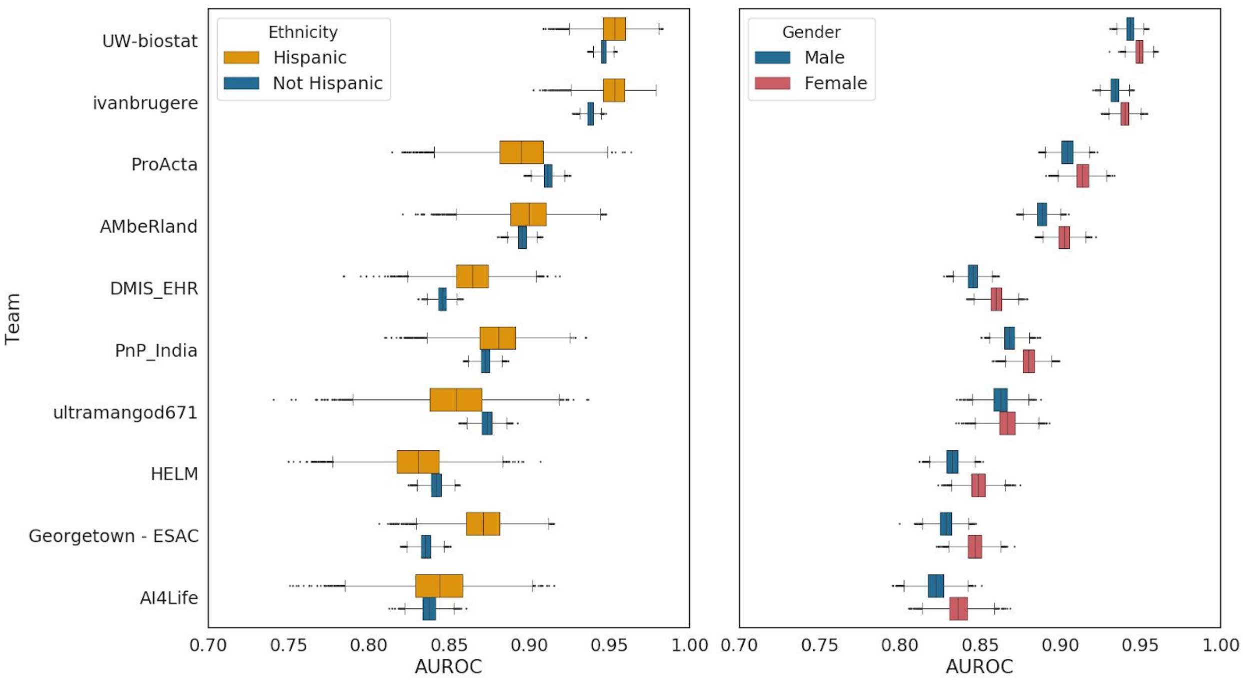


Supplemental Figure 4. Bootstrapped distributions (n=10,000) of the top 10 model AUROCs across two demographic variables: Ethnicity and Gender. Model predictions were randomly sampled with replacement and scored against the benchmark gold standard.

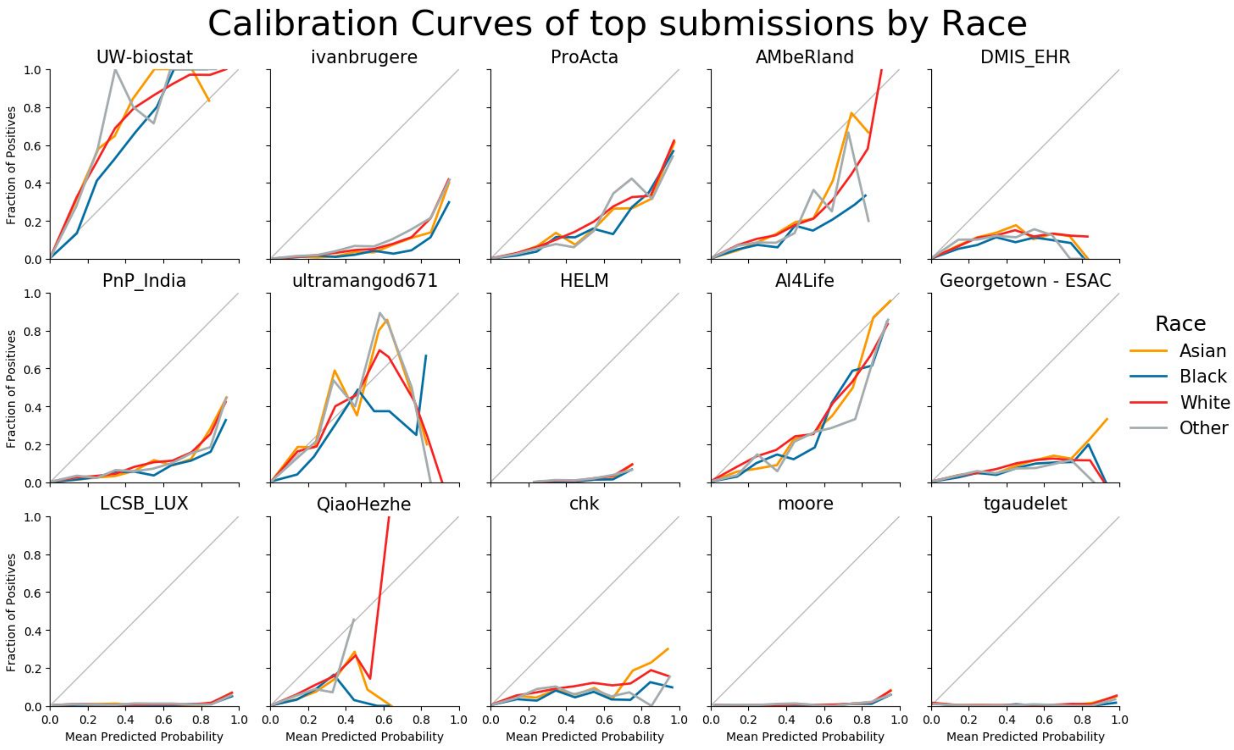


Supplemental Figure 5. Calibration curves from all final validation teams broken down by race.

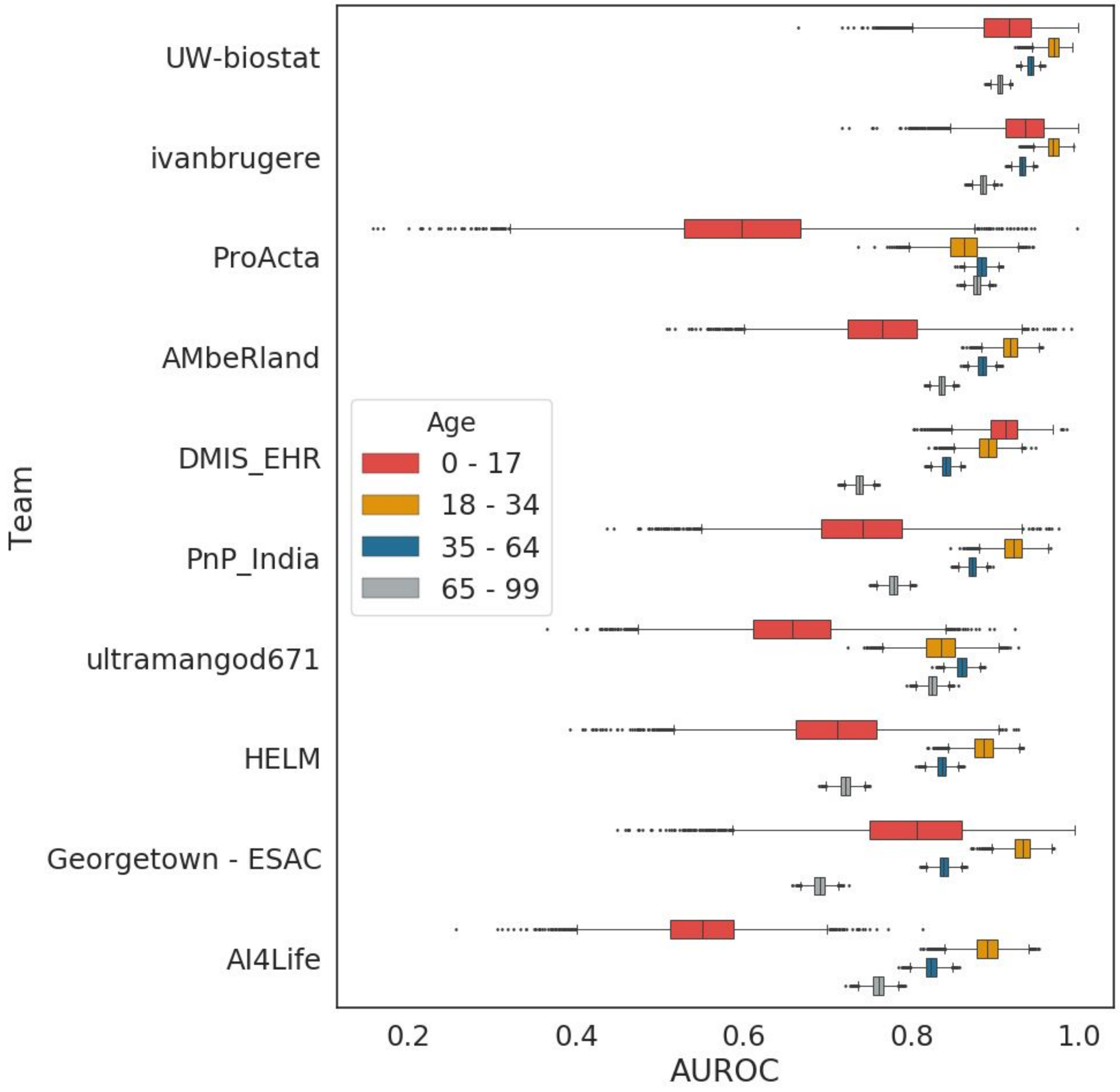


Supplemental Figure 6. Bootstrapped distributions (n=10,000) of model performance across age groups. The small number of patients age 0-17 results in high variance in model performance. For the most part, model performance is inversely correlated with age where models tend to me more accurate on 18-34 year olds and less accurate on 65-99 year olds.

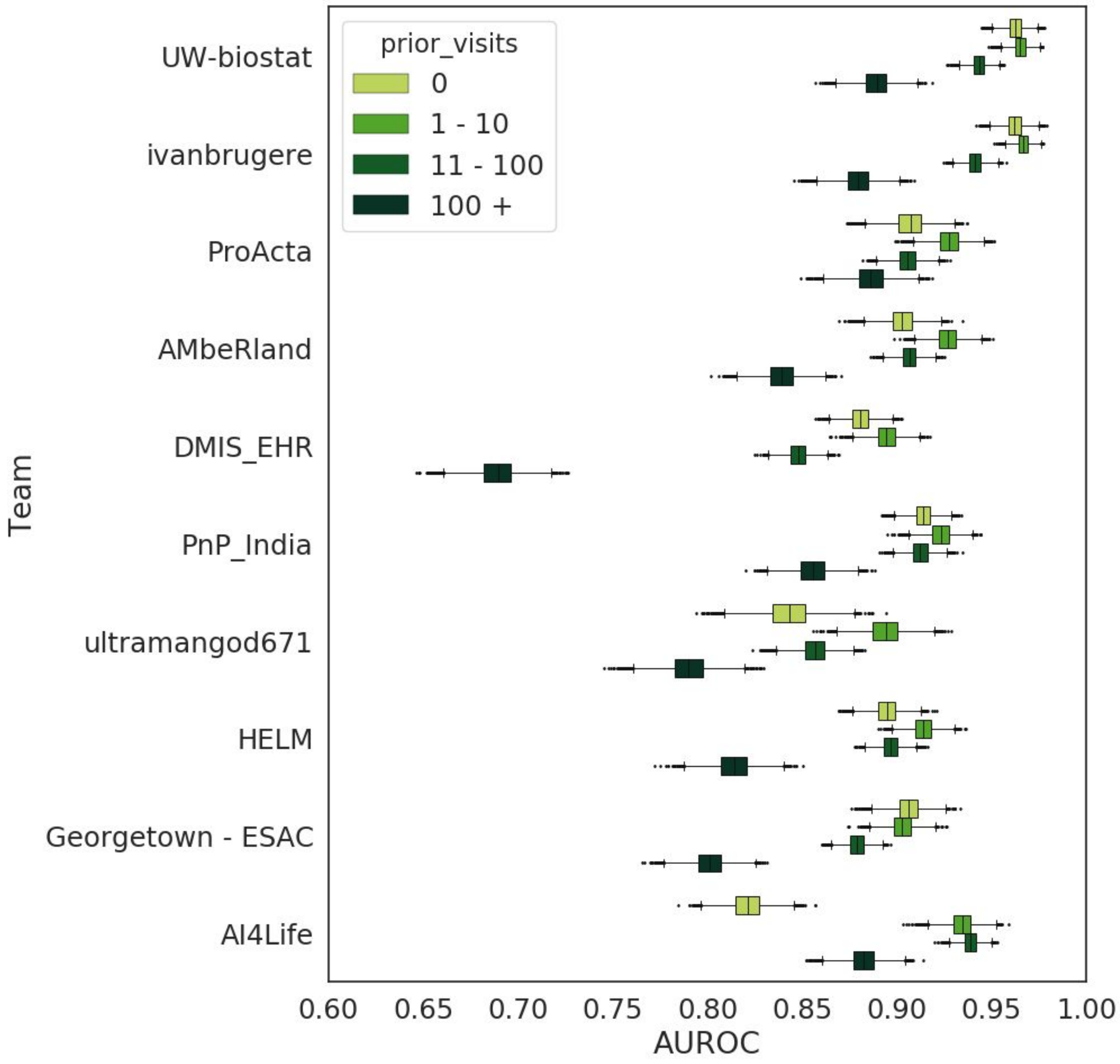


Supplemental Figure 7. Bootstrapped distributions (n=10,000) of model performance across the number of records available in each patient's clinical history. Most models perform the best on 1-10 records, with a decrease in accuracy on patient records with no history or 11-100 records, with a large decrease in performance on records of 100 or more.

#### Model Descriptions

We include the top 5 model descriptions from the validation phase. These models were not the most accurate during the leaderboard phase but were the highest performing models in the validation phase. We include these 5 descriptions as well as a table with links to the other submitted descriptions for all participating teams in the validation phase.

### UW-biostat

Jifan Gao[1,2], Guanhua Chen[1,2]
[1] Department of Biostatistics and Medical Informatics, University of Wisconsin-Madison, Madison, Wisconsin, USA
Contact: J.G., G.C.

#### Introduction

We use ontology-rollup to reduce the dimensionality and use time binning and sample reweighting to capture longitudinal characteristics. For the model part, we train and tune a LightGBM model to predict the mortality risk of each patient.

#### Methods

**Ontology-rollup**: We map each event in clinical findings (including diagnosis and symptoms), drug exposure, and procedure occurrence to medical terminology in ICD-9, ATC, and CPT, respectively. Such mapping information is available to download from the OHDSI and NIH websites.Then, by using the hierarchical structures of these vocabularies, we group together medical concepts which are clinically relevant. In particular, for clinical findings, their concept IDs are first mapped to an ICD-9 code and then mapped to the corresponding ICD-9 subgroup; for drug exposure, the drug IDs are first mapped to an ATC code and then mapped to the corresponding ATC pharmacological subgroup; for procedure occurrence, the procedure occurrence IDs are first mapped to a CPT code and then mapped to the corresponding CPT subgroup. As an example, any codes under ICD-9 section 317 (Mild intellectual disabilities), 318 (Other specified intellectual disabilities) and 319 (Unspecified intellectual disabilities) are grouped together as one concept to represent the condition of “Intellectual Disabilities”. In addition, we also count the ICD-9 codes of diseases that cause most deaths according to reports from the CDC (Center for Disease Control and Prevention) [1]. Such strategies lead to 459 features/mega concepts.

**Time binning**: To take into account the longitudinal data information in the EHR, we create five time frames with various lengths and summarize the occurrence of events within each time frame. Hence, there are five sets of these 459 mega concepts, one for each time frame. We also compute means and most recent values of selected measurements which are reported to indicate mortality [2]. Combining these features with other time-invariant information such as demographics, lifestyle, and disease history, a total of 2460 features are generated through this process. See Figure 1 for illustration.

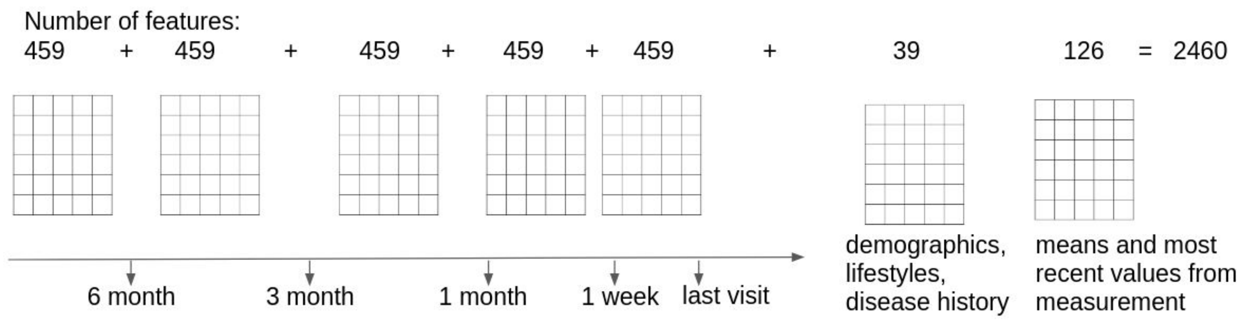


Figure 1. Feature Generation

**Missing label imputation:** As is described in the Challenge Website, some of the patients' mortality status (labels) are missing in the training data. Hence, there is a need to impute the missing labels. Among all strategies we have tried, the model produces the best AUROC results when all of these missing labels are set to be negative.

**Sample reweighting:** To take into account the potential time trend of mortality among patients in the EHR, we encourage our model to perform well for more recent patients. In particular, patients whose last visits are closer to the current date will be assigned a larger weight in the training process, whereas patients whose last visits are earlier will be assigned a smaller weight. The pseudo-labeled patients have weights of 1. The weights for those who are guaranteed to have a true label are computed as below:

$$weight = max(1, min(1.2, 1.2 - 0.2 \times \frac{JulianDay(\ last\ visit\ date\ ) - JulianDay(\ 2018 - 09 - 01, 12:00\ am\ )}{1000}))$$

Note that patients, whose last visit is before 2018-09-01 12:00 am, are guaranteed to have a true label. The function *JulianDay*(some DateTime) returns the DateTime as a Julian Day value, which is convenient for computation of DateTime in *YYYY-MM-DD HH:MM am/pm* Time format.

**Training/validation split:** In order to validate the model's "future-proof" ability, we order the labeled patients by their last visit date from recent to early and use the top 15% of patients for validation.

**Model Tuning:** We use Optuna [3] to enable automatic hyper-parameter tuning. AUROC is used as the stop criteria.

### Results

#### Model Performance

The AUROC and AUPR on the validation data are 0.9470 and 0.4779, respectively.

#### Feature Importance

The feature importance is computed by the LightGBM's built-in method, which is based on the gains of splits when a certain feature is used. The age of a patient at his/her last visit is the feature with highest importance. 7 of the top 10 features are related with measurement features such as pulse rates and blood pressures.

#### Discussion

Although our ontology-rollup strategy may lose variations of concepts within a group, the AUROC and AUPR results indicate that the overarching concepts can be represented by such a strategy. Compared with our latest model in the leaderboard phase, we add measurement features to our final model in the validation phase. Considering that the final model achieves highest AUROC and AUPR with the validation data, it is reasonable to believe our strategy of ontology-rollup, time binning and sample reweighting could potentially handle the data drift between the EHR datasets in the leaderboard phase and the validation phase. The importance of measurement features reflects that taking vital signs and lab results into more consideration may help to improve the performance of mortality risk prediction models.

#### Conclusion

We build a mortality risk prediction model based on LightGBM, with a dimensional reduction strategy tailored for sparse and longitudinal EHR data. The model achieves the highest AUROC and AUPR on the validation data, which are 0.9470 and 0.4779, respectively.

### IvanBrugere

#### Fine-tuning ensemble methods for tabular EHR data

Ivan Brugere[1], Lav Varshney[1]
[1]Salesforce Research
This submission is publicly available Submission code is available at: https://github.com/ivanbrugere/ehrdc/ (public after final leaderboard phase)

#### Background/Motivation

We focused on designing simple models with minimal feature engineering, model complexity, and hand coding. Initially, our focus was on sequential models which aimed to separate patient sequences into contiguous healthy and unhealthy segments. However, we found that static models outperformed these temporal models and allowed greater model exploration within the budget constraints of the competition.

A second principle we focused on was a common problem formulation which share pre-processing and featurization. This allowed for model selection over many models, and quick model development. Our final submission (syn21533058) achieves 0.9555 AUC without any domain-specific feature-engineering.

#### Methods

Our method is simple, minimizing the featurization and preprocessing needed for the prediction task. Our motivation was to show that very simple models would be effective on datasets an order of magnitude larger than prior EHR work. In this context, tree-based classifiers likely find very robust groups of discriminating features.

##### Preprocessing

We use the "OMOP useful columns" metadata as the subset of features we'll process. We construct a single [#patient x #features] matrix. All concepts are mapped to a boolean presence/absence rather than the real value, e.g. in the observation table. We treat age by unique years. In the table below, we summarize the number of unique concepts introduced in each table and field in the synthetic data (MD5: a488153640bfcc60d9c9a30e04c2169f):

| Table | Field | Unique Features (Concepts) |
|---|---|---|
| condition_occurrence | condition_concept_id | 3305 |
| procedure_occurrence | procedure_concept_id | 1376 |
| observation | observation_concept_id | 83 |
| measurement | measurement_concept_id | 7 |
| drug_exposure | drug_exposure_concept_id | 1293 |
| person | year_of_birth | 75 |
| person | gender_concept_id | 2 |
| person | race_concept_id | 6 |
| person | person_id | 97918 |

Note that person_id indexes the patient axis of our matrix, yielding a [97918 x 6147] patient x features boolean matrix on the included synthetic dataset. We process the death table conventionally, yielding a boolean vector with 1096 non-zero elements in the included synthetic dataset.

##### Models and Model Selection

We use repeated resampling of patient training rows to create equal-sized training/validation subsets. We refer to a method with its particular fixed hyperparameters as a "model." For each subset sample, we retrain each model and measure performance on the sampled validation subset. In the synthetic dataset (MD5: a488153640bfcc60d9c9a30e04c2169f) this yields a training and validation set both of size 48959.

Due to computational constraints, in our submission we limited our sampling to k=1 subsets so that we could evaluate more models. This, however, may not be good practice in a time-unconstrained setting. In our final submission, we found that catboost (https://catboost.ai/) had better performance through hyperparameter tuning than xgboost (https://xgboost.readthedocs.io/en/latest/) which, in turn, had better performance than scikitlearn traditional methods, deep learning embeddings, and sequential models. In our final submission (syn21533058), this yields a model selection over 12 catboost models with the following grid search:

| Hyperparameter | Values |
|---|---|
| depth | 6 |
| objective | log-loss |
| # trees | 3200 |
| learning rates | [0.01, 0.05, 0.075] |
| L2 leaf regularization | [10, 25] |
| class weights | [(0.1, 0.9), (0.25, 0.75)] |

Since we were unable to see which of these models is selected due to the privacy constraints, in prior submissions we also included xgboost baselines on some set of hyper-parameters. The intuition is that we can compete against current-best model in the model selection phase. We select the top-performing model with respect to mean AUROC on the validation-set samples, with no variance penalty.

#### Discussion: Other Methods

Through the development of our methodology, we experimented with many different models, which we briefly report. Although we have limited information on why these methods performed poorly, it may be illustrative and inspire further model/methodology development.

#### Tabular neural network methods

We learned supervised patient embeddings from input feature vectors, using varying network architectures. We included fully-connected architectures in model selection against prior best xgboost models. These models were trained over the following hyperparameters:

| Hyperparameter | Values |
|---|---|
| depth | [3, 4] |
| Layer1 | [200, 500, 50] |
| Layer2 | [100, 50, 20] |

| | |
|---|---|
| Layer3 | [50, 10] |
| Layer4 | [25, None] |
| learning rate | [0.01, 0.05] |

In addition to using the binary classification output layer, we trained base classifiers on (1) exclusively and (2) in addition to boolean input features. The performance of these models seemed to be bounded by the simpler xgboost base model on several submission tests, so we dedicated resources to investigating xgboost vs. catboost in the final submission.

#### Ensemble methods

We sampled some set of base models from our set of both neural network and xgboost classifiers to train an ensemble method which takes as input the model score outputs of the base classifiers and trains a final score output on labeled training data. For the ensemble model, we use a logistic regression with hyper-parameter tuning (LogisticRegressionCV in scikitlearn). In the constrained setting, we can train these models very quickly because we do not need to retrain the base models. However, we did not find a reliable improvement in performance after sampling several ($n$ = 20) of these models for differing number of base classifiers ($k$ = [4,6]) for a total of 40 sampled models.

#### Annual discriminator

We dedicated one submission to a slightly different prediction problem. These models discriminate between "healthy years" and "death years" by transforming patient rows to (patient, year) rows according to the same pre-processing pipeline. We use an xgboost classifier on the annual patient vectors. We return the non-zero model score only if the highest model score is the final year. We tested this model definition using the above model selection framework, to consider the final ($k$ = [1,2]) years as "death years," e.g. this classifier should distinguish the patient transitioning from healthy to unhealthy states, on annual time-steps.

It is plausible a more fine-grained sequential model either in terms of fixed time (e.g. semi-annual) or on visit sequences could discriminate this changepoint. However, we did not prioritize this line of work due to our low submission performance (syn21104894, AUC: 0.436).

#### Conclusion

We explored how simple gradient boosting models could be effective for large, tabular EHR datasets. However, there are several limitations of this work: First, due to the nature of the leaderboard evaluation, we could not qualitatively evaluate and iterate on our model with these findings. It is entirely possible that *uninformative* or spurious predictions are driving the model performance. Under the data use/privacy constraints of the challenge, further annotation of difficult examples may help control for strong covariates to mortality e.g. age. Second, we are unable to attribute the model performance to varying concept extraction regimes. In many instances, this featurization may itself be very difficult. Furthermore, there may be many hidden person-hours in coding, annotation which are not accounted for in model performance on raw data. Finally, since we are given this tabular featurization, our model is not designed for multimodal data such as clinical notes or medical imaging. Incorporating this data would be non-trivial though it could greatly improve interpretability and attribution.

#### Authors Statement

Ivan Brugere reviewed EHR related work, implemented all models, pre-processing, model selection methodology, and handled workflow/challenge troubleshooting. Lav Varshney provided input into modeling approaches investigated.

### Proacta

Zofia Nawalany[1] and Łukasz Charzewski[1,2]
[1]ProActa
[2]Division of Biophysics, Faculty of Physics, University of Warsaw
The submission will be made public as part of the challenge archive. Source code is available in the Synapse Project

#### Summary Sentence

We applied logistic regression on features reflecting frequency of medical events to predict the patients mortality in the next six months.

#### Introduction

We applied logistic regression (logit) [1] for a binary classification of patients based on their medical history. We decided to limit the considered data to the last 6 months of data available as the most recent data seems to have larger impact on the patients outcome.

#### Methods

##### Data Preparation

First, personal information were extracted from data and transformed accordingly. The age was divided by 100, gender and ethnicity were represented with binary and one-hot encoding, respectively.
Patients medical history was represented in form of events frequency in a given time:

- Visits table were employed to construct three features based on visit_concept_id. Each occurrence were counted with sklearn's [2] CountVectorizer.
- Useful concept IDs form procedures, conditions and observations in each patient were counted, also with CountVectorizer.
- Drug exposure entries were translated to active substances using Athena database, counted with MultiLabelBinarizer.
- Useful concept IDs of measurements were counted with MultiLabelBinarizer.
- Created values were directed to feature selection (with SelectKBest utilizing chi2 scoring) retaining 50 features where possible.

Submissions with dimensionality reduction (PCA preceded with StandardScaler) instead of KBest selection were tried as well, but they performed worse, especially due to AUPR metric. Also oversampling the underrepresented class with SMOTE algorithm was tested, but rejected, as the AUROC metric decreased. To assure that patients with death were not registered didn't die indeed, their data were truncated for 180 days before the last available entry. In such prepared data only the last 180 days of patients history were considered.

##### Model

Logistic regression [1] model was chosen due to its confirmed, diverse applications in biology and medicine. Also, its direct ability to predict the probability of class affiliation is desired for AUROC and AUPR metrics. Model fitting was performed with default sklearn's parameters.

#### Conclusion/Discussion

The obtained results with out model are satisfying. It is worth noting though, that some features used don't describe patient condition per se. The way of measurements processing reflects rather medical stuff opinions and decisions. Utilization of measurements results values would be needed for better patients description, but these may require detailed insight in their potential values.


#### Authors Statement

Both authors contributed equally in model design, data preparation and analysis.

### AMbeRland

Renata Retkute[1], Alidivinas Prusokas[2], Augustinas Prusokas[3]

[1] Department of Plant Sciences, University of Cambridge, Downing Street, Cambridge, UK

[2] School of Natural and Environmental Sciences, Newcastle University, Newcastle, UK

[3] Department of Life Sciences, Imperial College London, London, UK

#### Introduction

Gradient tree boosting was utilised to predict the probability of mortality from Electronic Health Records. This method combines boosting and optimisation to allow accurate predictions. We modeled this using the *gbm* package in R [1], which implements AdaBoost's exponential loss function (its bound on misclassification rate) and uses Friedman's gradient descent algorithm [2]. We utilised this algorithm in two stages - an initial and final model. The final model showed high predictive performance: the AUROC for leaderboard phase was 0.9252 and for validation phase was 0.8954.

#### Methods

We predicted mortality based on the presence or absence of certain features in the personal records of each patient.

First, the initial model was trained on features marked present in patients having status "death". The relative influence of each feature in the model was calculated, and features with a relative influence larger than zero were retained. The final model was then trained on these filtered features, to generate a simplified but just as effective model. A diagram of stage 1 is shown below.

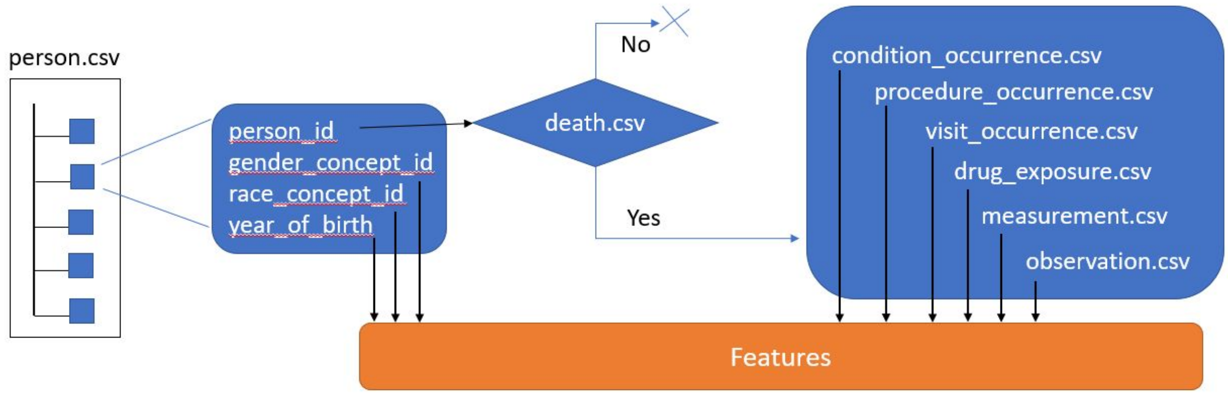


Features for stage one were selected as follows:

- 'gender_concept_id', 'year_of_birth' ,'race_concept_id';
- conditions from "condition_occurrence.csv" dataset which were present in at least one patient with status="death"; conditions were coded 0 for absence and 1 for presence;
- procedures from "procedure_occurrence.csv" dataset which were present in at least one patient with status="death"; procedures were coded 0 for absence and 1 for presence;
- drug exposure from "drug_exposure.csv" dataset which were present in at least one patient with status="death"; drug exposure were coded 0 for absence and 1 for presence;
- measurements from "measurement.csv" dataset which were present in at least one patient with status="death"; measurements were coded 0 for absence and 1 for presence;
- observations from "observation.csv" dataset which were present in at least one patient with status="death"; observations were coded 0 for absence and 1 for presence;
  -visit occurence from "visit_occurrence.csv" dataset which were present in at least one patient with status="death"; visit occurrence were coded 0 for absence and 1 for presence.

The following options were used to train both Generalized Boosted Regression Models:

- distribution = "bernoulli",
- n.trees = 100,
- interaction.depth = 1,
- cv.folds = 5.

### Results

#### Model Performance

In the overall leaderboard phase, our mortality prediction model was ranked 12$^{th}$ (AUROC=0.9252, AUPR=0.1672). In the validation phase, it was ranked 4$^{th}$ (AUROC=0.8954, AUPR=0.1656). Therefore, the model performed well on data accumulated in the UW environment before February 2019 (leaderboard phase), and on data accumulated from February 2019 to January 2020 (validation phase).

#### Feature Importance

As features were selected adaptively, we received feature evaluation after the challenge was closed.

### Discussion

Gradient tree boosting has been previously used for predicting cardiovascular risk [3], mortality in a large general oncology population [4], and heart failure outcomes [5]. Our submission had the AUROC for leaderboard phase 0.9252 and for validation phase 0.8954, showing that gradient tree boosting can achieve high accuracy in prediction of mortality. Furthermore, this relatively low drop in AUROC between the different data domains (pre- and post- February 2019) shows a high generalisation and robustness in the algorithm, possibly due to its simplification by the removal of unnecessary features.

Unlike previously developed automatic mortality prediction models, the EHR DREAM Challenge offers a different philosophy for ML: model improvement is driven more by intuition and domain knowledge than by precise analysis of the model, as the developer has access only to synthetic data.

### Conclusion

ML-based prognostic tools can help to develop a holistic view of patients' health, but they face the challenge of a huge amount of data and complex pipeline selection and configuration. We have proposed a conceptual algorithm which is straightforward to implement, generalises well, and does not require much data pre-processing or high computation cost in terms of model training and inference.

### DMIS_EHR

#### Patient Mortality Prediction with Concept Record Embedding

Yonghwa Choi[1], Sanghoon Lee[1], JunSeok Choe[1], Inggeol Lee[2], Sunkyu Kim[1] and Jaewoo Kang[1,2]
[1] Department of Computer Science and Engineering, Korea University, Seoul, Korea
[2] Interdisciplinary Graduate Program in Bioinformatics, Korea University, Seoul, Korea

#### Summary Sentence

We mapped each concept ids to concept embedding space using 'Word2Vec' algorithm, and used gradient boosting based algorithm with concept embedding vectors to predict patients' mortality.

#### Background/Introduction

In EHR dataset, each patient has several visit records, and each visit has several feature concepts. We consider these feature concepts that appear for each visit as a word token and the visit occurrence as a sentence - a set of word tokens. If we consider such records as words and sentences, we can apply NLP techniques and algorithms, especially word embedding. Word embedding is one of the most popular feature learning techniques in natural language processing. Each word is mapped to a real number vector which has a much lower dimension than that of word token. The most common embedding algorithm is word2vec. The word2vec algorithm takes lots of sentences as an input and generates word embedding of sentences in specific embedding dimensions. The generated embedding vector can be used to predict the mortality of patients through several classification models. Gradient boosting is one of the most powerful machine learning algorithms, which generates a model in the form of an ensemble of weak decision trees. However, as the provided dataset is too large to apply normal gradient boosting algorithm, LightGBM can be a good option, which is designed to be used with faster training speed and better performances.

### Methods

#### Data preprocessing

Feature: Initially, we parse 5 feature files: condition_occurrence.csv, drug_exposure.csv, observation.csv, measurement.csv and procedure_occurrence.csv. We extract visit_occurrence_id and concept_id from each line of the file, and the list of concepts is mapped to the corresponding visit. These concepts are considered as set of word tokens, so it can be used to generate concept embedding through the word2vec algorithm. The order of words in a sentence is a key factor in capturing relationships between words. However, since the data we process has no order, we experimented with changing the window size of the word2vec algorithm, but there was no performance difference with the synthetic data.
Demographic: From the “person.csv” file, we extract person_id and several other demographic features such as year, race, and gender, with the default label as “False.” After reading all patients’ information, we calculated the average year of birth of all patients for imputation, as we found some of the patients had no corresponding data. As for the rest of the demographic features, we used them as binary features.
Visit occurrence: While reading the “visit_occurrence.csv” file, we store a list of visit_occurrence_ids and its date to track when the person visited. In the training phase, we read the “death.csv” file to track who died within the 6 months after the last visit, and set the label as “True.” In the validation phase, labels of people in the validation dataset stay as “False,” because we only use the output of the prediction.

#### Model

After preprocessing, we generate patient representations using concept embedding and demographic features. We calculate the concept representation as a sum of the embeddings of all unique concepts for each patient and concatenate demographic features to it. When we use time interval decay in visits, concept representation is calculated as a sum of weighted concept embeddings with their interval decay values as weights. We’ve experimented with various time decay methods, but the performance was rather poor when we submitted them to the leaderboard lane. Then the generated patient representations are passed to the classification model. When we tested GradBoost and LightGBM for the classification model, LightGBM was much faster and showed comparable or even better performances in validation situations.

#### Experiment

20% of the training data, called “valid” data, was split, keeping a similar distribution with the rest of the data to evaluate our approach and parameters. Models are trained with the rest of the data and its parameters are chosen according to the performance test in the “valid” data. The candidates for the parameters were chosen as the ones that performed well in the local environment with the synthetic dataset. In the leaderboard submission, however, training with all data and the parameters selected with the synthetic dataset without cross-validation performed the best, and its parameters are listed below.

| Parameter name | Value |
|---|---|
| learning_rate | 0.007955749 |
| n_estimators | 3734 |
| reg_alpha | 0.992386407 |
| reg_lambda | 0.006758808 |

#### Conclusion/Discussion

We propose a clinical concept embedding method based on considering EHR visit records as sentences and each record concept as a word token using only concept ids and visit occurrences. In order to capture the relationship between EHR record concepts, an embedding method is shown to be a good option to predict the mortality of patients, although the model used to predict is quite small and simple. Using deep learning algorithms, such as Recurrent Neural Networks or attention mechanisms, and external knowledge, would improve the performance.


All teams implemented a form of dimension reduction to operate in a far smaller modeling space than the 1.2 million unique granular concepts available in the UW dataset. Team UW-biostat derived 1405 weighted features from 1.2 million unique granular concepts; Team IvanBrugere used 1479 weighted features representing 1479 unique granular concepts; Team Proacta used 224 weighted features representing 22,934 unique granular concepts; Team AMbeRand used 35 weighted features representing 35 unique granular concepts; and Team DMIS's model used a Principal Component Analysis to generate principal components as features for their trained model.

Supplemental Table 3. A list of the final 15 model write ups that were submitted to the validation phase for evaluation.For the top 4 models, we report the features and unique concepts used in the model during the validation phase of the challenge.

| Final Rank | Team | Project URL | Method | Number of Weighted Features | Unique concepts |
|---|---|---|---|---|---|
| 1 | UW-biostat | https://www.synapse.org/#!Synapse:syn21598356/wiki/601151 | LightGBM | 1,405 | 1.2 million |
| 2 | ivanbrugere | https://www.synapse.org/#!Synapse:syn20833371/wiki/600725 | CatBoost | 1,479 | 1,479 |
| 3 | ProActa | https://www.synapse.org/#!Synapse:syn20927255/wiki/601202 | Logistic Regression | 224 | 22,934 |
| 4 | AMbeRland | https://www.synapse.org/#!Synapse:syn21301805/wiki/601083 | Generalized Boosted Regression | 35 | 35 |
| 5 | DMIS_EHR | https://www.synapse.org/#!Synapse:syn21569020/wiki/600891 | LightGBM | | |
| 6 | PnP_India | https://www.synapse.org/#!Synapse:syn21962250/wiki/602171 | XGBoost | | |
| 7 | @ultramangod671 | https://www.synapse.org/#!Synapse:syn21962254/wiki/602172 | LightGBM | | |
| 8 | HELM | https://www.synapse.org/#!Synapse:syn21962268/wiki/602173 | XGBoost | | |
| 9 | AI4Life | https://www.synapse.org/#!Synapse:syn20838956/wiki/601297 | XGBoost | | |
| 10 | Georgetown - ESAC | https://www.synapse.org/#!Synapse:syn21624375/wiki/601326 | XGBoost | | |
| 11 | LCSB_LUX | https://www.synapse.org/#!Synapse:syn21962299/wiki/602176 | Random Forest | | |
| 12 | @QiaoHezhe | https://www.synapse.org/#!Synapse:syn21962317/wiki/602177 | XGBoost | | |
| 13 | @chk | https://www.synapse.org/#!Synapse:syn21962336/wiki/602178 | Logistic Regression | | |
| 14 | moore | https://www.synapse.org/#!Synapse:syn21962343/wiki/602179 | XGBoost | | |
| 15 | @tgaudelet | https://www.synapse.org/#!Synapse:syn21962356/wiki/602181 | Neural Network | | |
